# FedWeight: Mitigating Covariate Shift of Federated Learning on Electronic Health Records Data through Patients Re-weighting

**DOI:** 10.1101/2025.02.10.25322018

**Authors:** He Zhu, Jun Bai, Na Li, Xiaoxiao Li, Dianbo Liu, David Buckeridge, Yue Li

## Abstract

Federated learning (FL) enables collaborative analysis of decentralized medical data while preserving patient privacy. However, the covariate shift from demographic and clinical differences can reduce model generalizability. We propose FedWeight, a novel FL framework that mitigates covariate shift by reweighting patient data from the source sites using density estimators, allowing the trained model to better align with the distribution of the target site. To support unsupervised applications, we introduce FedWeight ETM, a federated embedded topic model. We evaluated FedWeight in cross-site FL on the eICU dataset and cross-dataset FL between eICU and MIMIC III. FedWeight consistently outperforms standard FL baselines in predicting ICU mortality, ventilator use, sepsis diagnosis, and length of stay. SHAP-based interpretation and ETM-based topic modeling reveal improved identification of clinically relevant characteristics and disease topics associated with ICU readmission.

## Introduction

Training machine learning (ML) models on large-scale electronic health record (EHR) data is promising for advancing medical research and improving patient outcomes [1]. However, EHR data are from different healthcare institutions are not easily pooled, because it is difficult to move these data out of institutions due to laws and regulations about data privacy and data governance, and transmission costs [2]. To enable machine learning in the context of these challenges, Federated Learning (FL) trains the model on local datasets that remain in each healthcare institution and shares only the model parameters with the central server for model averaging [3]. Although FL has been widely used in the clinical settings [4–6], traditional FL assumes the same data distributions for each silo [7–12]. This is an unrealistic assumption because of covariate shifts due to differences in patient demographics, clinical practices, and data collection methods between institutions [13, 14]. Such disparities may lead to poor performance on out-of-distribution (OOD) clinical data, resulting in inaccurate predictions and uninterpretable clinical outcomes. For instance, a federated model trained on data from specialized hospitals may be unable to accurately predict patient outcomes at a community hospital. Beyond common supervised tasks in FL affected by covariate shifts, unsupervised tasks, such as topic modeling techniques, like Embedded Topic Models (ETM) [15] for extracting latent representations from high-dimensional EHR data, are also vulnerable as ETM’s topic distributions can vary significantly across institutions. Therefore, given the significance of AI safety and quality [16–19], it is essential to develop a framework to mitigate the effect of covariate shifts on FL, thus providing more robust inference to enhance disease prevention strategies and promote fairness in healthcare decisions and outcomes [20–22]. In this study, we aim to develop an FL-powered medical modeling framework for EHR data that mitigates covariate shifts by re-weighting patients from source clinical sites to align with the target site’s data distribution, thereby improving model generalization and clinical outcome predictions.

Recently, Shimodaira *et al*. introduce the weighted log-likelihood method to address covariate shift using importance sampling, assigning weights to the training loss based on the ratio of test to training input densities [13]. Building upon their work, methods have been proposed to estimate the reweighting ratios using Kernel Mean Matching [23, 24], as well as Kullback-Leibler Importance Estimation Procedure (KLIEP) [25], which have been extensively employed to alleviate covariate shifts in centralized learning environments. However, all these methods rely on access to both training and test samples of all patients to estimate the reweighting ratios, which is impractical in FL settings due to privacy constraints. To mitigate this challenge, FedCL and FedDNA employ a model-level reweighting approach by sharing statistical model parameters across clients [26, 27], but their improvements are limited as the reweighting is estimated solely at the model level. In contrast, several existing methods estimate the sample-level reweights by sharing unlabelled data across clients [28, 29], which may increase communication costs and compromise data privacy, thus violating FL principles. To address this issue, FedDisk trains a global density estimator along with multiple local density estimators to compute reweighting ratios [30], but their experiments demonstrate only minor improvements on real-world image data. Moreover, other enhanced FL methods such as FedProx [31], SCAFFOLD [32], MOON [33], and FedNova [34] address general data heterogeneity by introducing regularization, control variates, or normalization-based aggregation. For instance, FedProx adds a proximal term to the local objective to constrain divergence from the global model. While these methods improve training stability under data heterogeneity, they primarily mitigate optimization variance and do not explicitly address covariate shift—a critical issue in clinical settings where input distributions vary across institutions (e.g., due to demographic differences). Furthermore, their lack of sample-level importance weighting limits their ability to correct distribution mismatches between training and deployment environments. In summary, existing methods either provide limited performance improvements or compromise privacy, making them unsuitable for healthcare, where patient confidentiality is paramount. Given that FL in healthcare is still nascent, with limited algorithms addressing covariate shifts [35, 36], there is a pressing need to develop a privacy-preserving solution with improved performance in clinical settings.

In this study, we first describe covariate shifts in two widely used public EHR datasets, namely the eICU Collaborative Research Database (eICU) [37] and the Medical Information Mart for Intensive Care III (MIMIC-III) [38]. We then present a novel FL framework — Federated Weighted Log-likelihood (FedWeight). Specifically, FedWeight incorporates the weighted log-likelihood method within the federated framework [13], which probabilistically re-weights the patients from the source clinical sites, aligning the trained model with the data distribution of the target site. FedWeight can be applied to both supervised and unsupervised tasks. We evaluate our framework using eICU and MIMIC-III datasets. Within the eICU, we perform cross-hospital FL. We then conduct federated training between eICU and MIMIC-III, while addressing their distribution differences. The experiments demonstrate that compared with the existing methods, our approach can provide more accurate predictions of patient mortality, ventilator use, sepsis diagnosis, and ICU length of stay.

## Results

### Identifying covariate shifts in clinical data

#### ML-based data harmonization

Different hospitals may adopt distinct administration practices, leading to variations in naming the same drug across clinical sites. Specifically, in the eICU dataset, different hospitals may use distinct encoding of drug administration. Some may use the generic name (e.g. “acetylsalicylic acid”), while others use the trade name (e.g. “aspirin”), although both refer to the same drug. Moreover, the dosage information is included in some drug names (e.g. “aspirin 10 mg”) but not all. As a result, we observed distinct clusters of patients by hospitals (Figure 1a). Furthermore, over 40% of drug names are not recorded (Figure 1d), although some of them have Hierarchical Ingredient Code List (HICL) codes. However, the HICL codes are not widely used in other hospitals, such as the one in MIMIC-III, which impedes the training of FL models across hospitals and datasets. In addition, there is almost no overlap in drug encodings across hospitals (Figure 1f). To address this, we developed a method to impute missing drug names. We also developed a drug harmonization framework to combine drugs with similar identities and exclude dosage information (see “Data preprocessing” in **Methods**). This preprocessing step successfully decreased the unrecorded drug proportions to approximately 20% (Figure 1e) and increased the number of common drugs to over 90%. The clustering of patients from different hospitals shows better mixing, although some hospitals still exhibit distinct clusters (Figure 1b). To prepare for model training across datasets, we also harmonized data between two data domains namely eICU and MIMIC-III (Figure 1c). Interestingly, we observed two distinct clusters from the MIMIC-III drug data (Supplementary Figure 1a). Enrichment analysis revealed that one cluster is significantly associated with planned hospital admissions (elective ICU admission) and cardiovascular surgery patients (Supplementary Figure 1b, c). These covariate shifts due to the heterogeneous distributions between hospitals and between datasets motivate us to develop FedWeight as experimented next.

**Figure 1:**
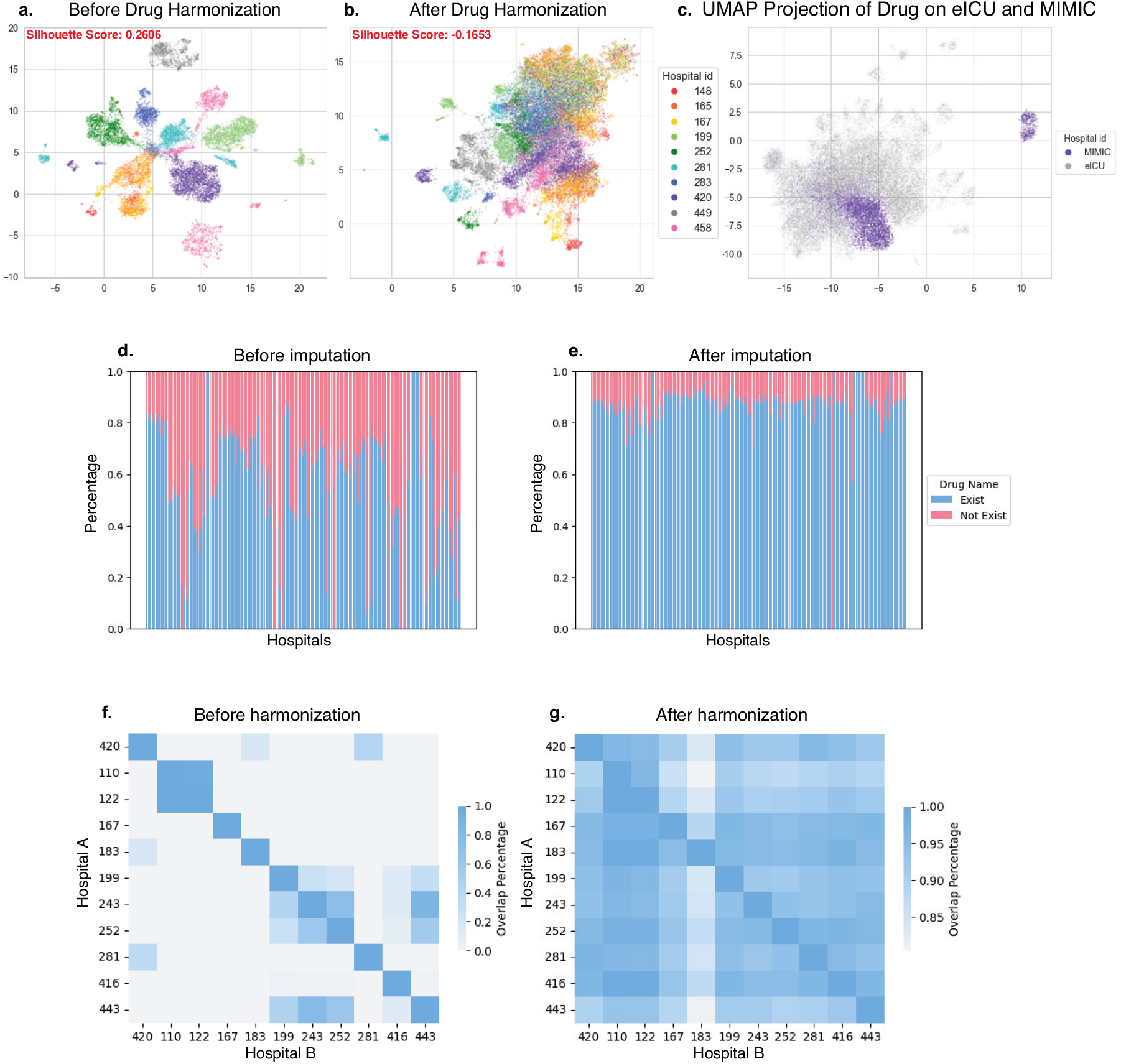
Effect of drug imputation and drug harmonization in the eICU dataset. **a** - **b**. UMAP projection of drug data across hospitals on eICU before and after drug harmonization. The UMAP-reduced drug data is visualized in a scatter plot, with each data point coloured by its respective hospital. The Silhouette Score [39] measures the degree of mixing, where a lower score indicates better mixing of drug data across hospitals. **c**. UMAP projection of harmonized medication data from eICU and MIMIC-III. **d** - **e**. The drug rates before and after imputation of the eICU data. Since some drugs may lack recorded names, drug rates indicate the proportion of administered drugs with recorded names over all unique drugs. **f** - **g**. Percentage of overlapped drugs before and after harmonization across the eICU hospitals. The percentage was computed as the proportion of drugs with recorded names present in both Hospital A and B relative to all drugs in Hospital B.

#### Heterogeneous patient demographics across eICU hospitals

In addition to the drug data, we also observed demographic differences among the hospitals in the eICU dataset. Specifically, we analyzed the distribution of patients by age, sex, BMI, and ethnicity within each hospital (Figure 2a-d). Although most patients were between 50 and 89 years old, Hospital 148 had a higher proportion of younger patients (< 30) (Figure 2a). In addition, Hospital 458 had a higher proportion of ethnically African patients, whereas Hospitals 167 and 165 had more Native American patients (Figure 2b). Despite the overall low proportion of underweight patients across all hospitals, Hospital 199 manifested a slightly lower proportion of underweight patients and a higher proportion of obese patients (Figure 2c). Furthermore, while most hospitals had more male patients than female patients, Hospital 283 had an almost equal proportion of male and female patients (Figure 2d). These discrepancies in the demographics of patients in hospitals can hinder the effective generalization of models in FL settings.

**Figure 2:**
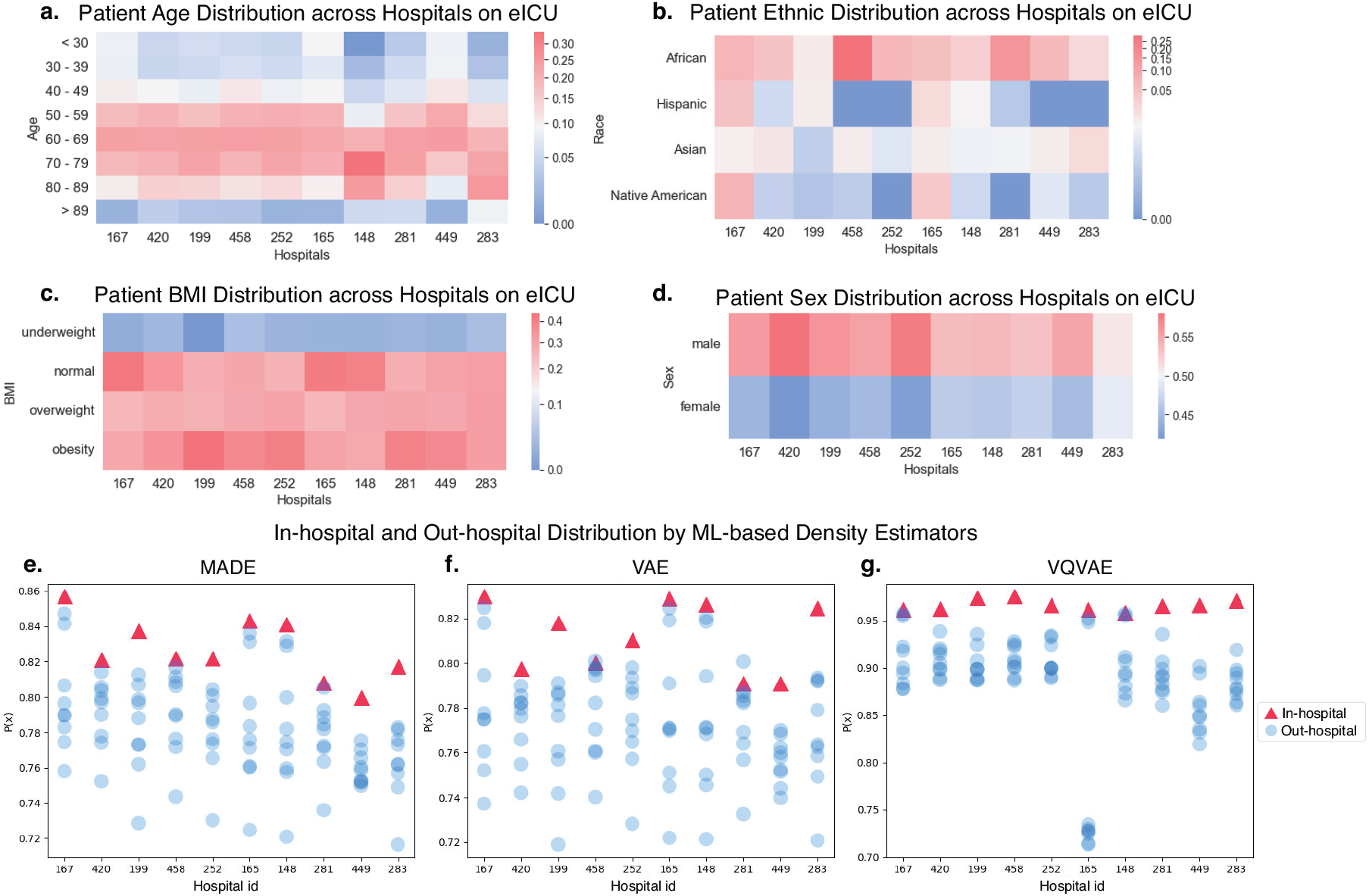
Visualization of covariate shift in the eICU and MIMIC-III dataset. **a - d**. Distribution differences in patient demographics, calculated as the proportion of patients in each demographic group within each hospital. Note: The Caucasian group was excluded from panel b to enhance the visibility of minority group distributions, as its majority presence would otherwise dominate the color scale and mask smaller variations. **e** - **g**. In-hospital and out-hospital distribution by ML-based density estimators. For each hospital, the in-hospital estimate was calculated by applying its own density estimator to its patient data. The out-hospital estimate was computed by applying the same density estimator to patient data from a different hospital.

#### Quantifying data likelihood by ML-based density estimators

To address the problem of covariate shift, we first measured data distributions using model-based approach. To this end, we experimented with 3 deep learning density estimators, namely Masked Autoencoder for Density Estimation (MADE) [40], Variational Autoencoder (VAE) [41], and Vector Quantized Variational Autoencoder (VQ-VAE) [42]. Through each model, we can compare the data likelihoods between the training data from one hospital and test data from another hospital. In most hospitals, the in-hospital likelihood is significantly larger than the out-hospital likelihood (Figure 2e-g), indicating that each hospital’s model aligns more closely with its own data than other hospitals’ data. The results also show that the 3 density estimators possess the sensitivity to detect covariate shifts.

### Addressing covariate shifts by sample re-weighting

Putting the above together, we developed FedWeight, a novel model-agnostic ML method to re-weight the patients from source hospitals, aligning the trained models from each source with the data distribution observed in the target hospital (Figure 3a-d). Specifically, the target hospital shares its density estimator with all source hospitals, which use it to calculate the reweighting ratio to train their local models. Intuitively, we assign larger weights to the patients from the source hospitals whose data distributions similar to the target population and smaller weights to those with dissimilar data distributions. Consequently, the trained model is better aligned with the target hospital data distribution, thereby effectively addressing the covariate shift problem in the FL settings. During training, only model parameters are shared between the target hospital and the source hospitals, thus effectively safeguarding patient privacy.

**Figure 3:**
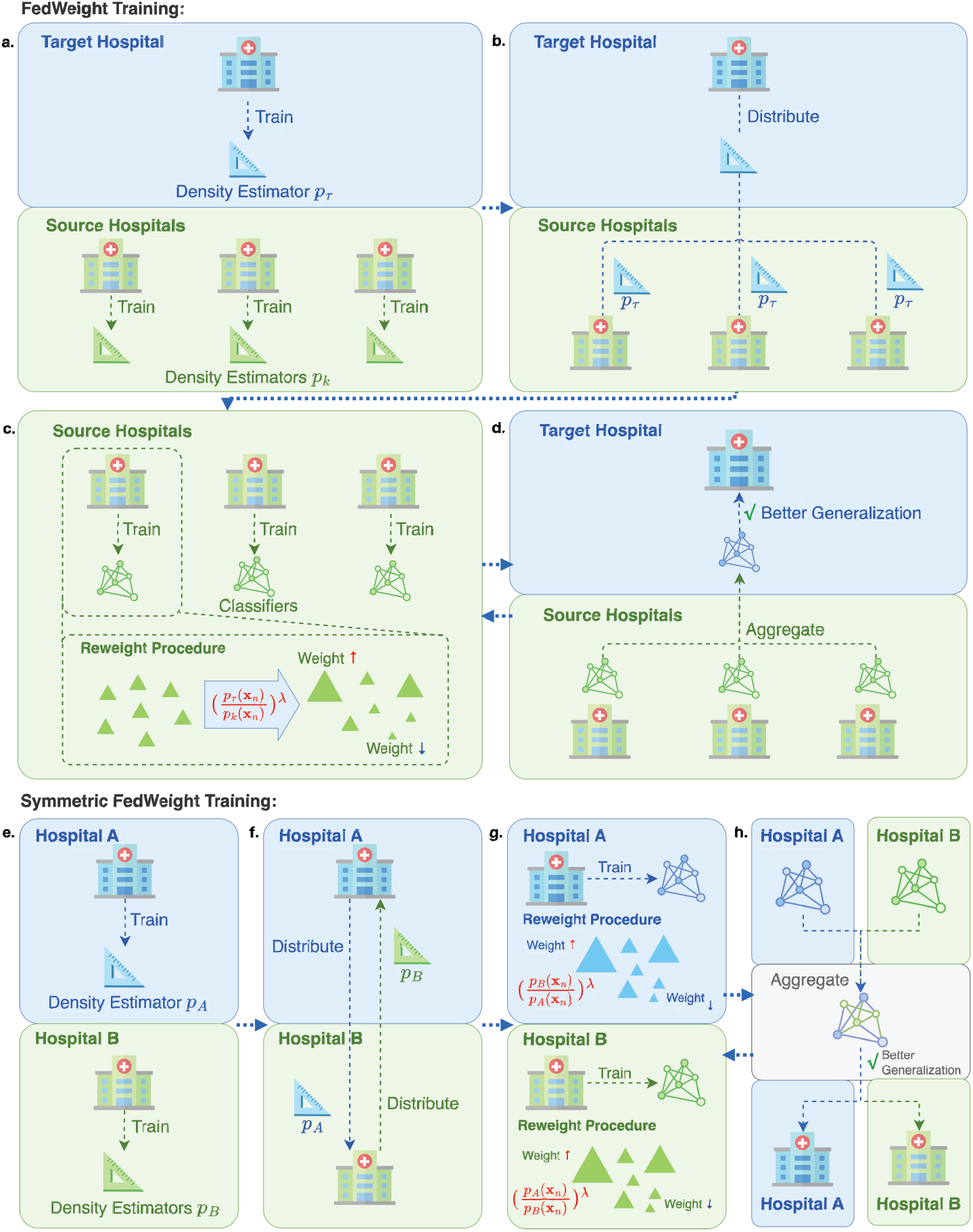
FedWeight training process. **a**. The target and source hospitals independently train their density estimators. **b**. The target hospital shares its density estimator with all source hospitals. **c**. Each source hospital calculates a patient-specific re-weight, which is used to train its local model. **d**. The target hospital aggregates the parameters from the source hospitals using the FL algorithm, which better generalizes the target hospital’s data. Symmetric FedWeight training process. **e**. Hospital A and B independently train their density estimators. **f**. Hospital A and B share their density estimators to each other. **g**. Hospital A and B regard each other as targets, estimating the re-weight, which is used to train its local model. **h**. Aggregate the local models from Hospital A and B using FL algorithm.

This method facilitates adapting source hospital data to the target. Additionally, we could also allow hospitals to mutually benefit from each other. To achieve this, we developed Symmetric FedWeight (Figure 3e-h). For simplicity, we assumed a federated network with two hospitals, which can be easily extended to multiple hospitals. These hospitals will treat each other as the target and themselves as the source to calculate the reweighting ratio (Figure 3g). The symmetric strategy enables both hospitals to adapt to each other’s data distributions.

### Simulation study

To evaluate the effectiveness of our method, we first undertook simulation experiments on patients’ data. We trained the FedWeight model on the simulation dataset by employing density estimators, namely MADE, VAE, and VQ-VAE. Our simulation mimics the label imbalance in the real-world data (e.g., more disease cases than healthy controls). To prioritize precision over recall, we used the Area Under the Precision-Recall Curve (AUPRC) as the evaluation metric for model prediction. To assess the statistical significance of AUPRC values between FedWeight methods and the baseline FedAvg, we performed Wilcoxon rank-sum test (also known as the Mann–Whitney U test) [43], a non-parametric test that is suitable for the limited number of AUPRC values in our federated model scenario [44]. We observed that all FedWeight models achieve an average AUPRC of 0.923, which significantly surpassed that of FedAvg, which had an average AUPRC of 0.917 (Wilcoxon rank-sum test p-value *<* 0.05) (Supplementary Figure 2a). This result demonstrates that our proposed model has enhanced predictive capability over this baseline with the datasets examined.

We also examined FedWeight’s capability in identifying influential features. Specifically, we evaluated the trained model by comparing its weights to a sparse reference model that simulates true influential features, using AUPRC as the evaluation metric. FedWeight models, when trained with density estimators such as VAE, VQ-VAE, and MADE, achieve comparable AUPRC results for detecting influential features (Supplementary Figure 2b). All FedWeight models significantly outperform the baseline FedAvg, demonstrating their potentials to pinpoint important biomarkers for real-world applications (see “Detecting clinical features by FedWeight+SHAP analysis”).

Additionally, we computed the Pearson correlation between the weights of the reference model and the trained model. We observed that FedWeight exhibits a higher correlation with the reference model’s weights (Supplementary Figure 2c). Therefore, FedWeight is more effective in capturing influential features, thus providing better model interpretability.

### Predicting critical outcomes from eICU data

Accurately predicting critical clinical events can drastically improve patient outcomes, especially in the ICU. Inspired by prior studies [45–48], we conducted experiments to predict four outcomes, namely mortality, ventilator use, sepsis, and ICU length of stay, using the first 48 hours of data from the eICU dataset (see “Data preprocessing” in **Methods**). These outcomes were selected based on their clinical importance, task diversity, and prevalence in existing FL research. Each plays a key role in ICU care: mortality prediction supports early risk stratification; ventilator use and sepsis prediction enable timely intervention and resource planning; and ICU length of stay aids in discharge management and capacity planning. Moreover, the four outcomes cover both classification (mortality, ventilator use, sepsis) and regression (length of stay) tasks, as well as both fixed-point (mortality, length of stay) and sequential (ventilator use, sepsis) prediction settings. This diversity enables a comprehensive evaluation of model performance across varying clinical and algorithmic scenarios.

To demonstrate the benefits of FL, we first compared the performance of non-FL (i.e., training on one hospital and tested on another) and FL methods. Our results show that all FL methods outperformed the non-FL method (Supplementary Table 1). Then, we compared the performance of FedWeight with both FedAvg and FedProx. Overall, FedWeight consistantly outperformed FedAvg across most hospitals and demonstrated competitive or superior performance compared to FedProx (Figure 4a-d; Supplementary Table 2). Additionally, FedWeight variants also demonstrated performance close to that of the centralized model, which was trained on pooled of data from all hospitals using hospital IDs as one of the covariates to account for hospital-specific batch effects.

**Figure 4:**
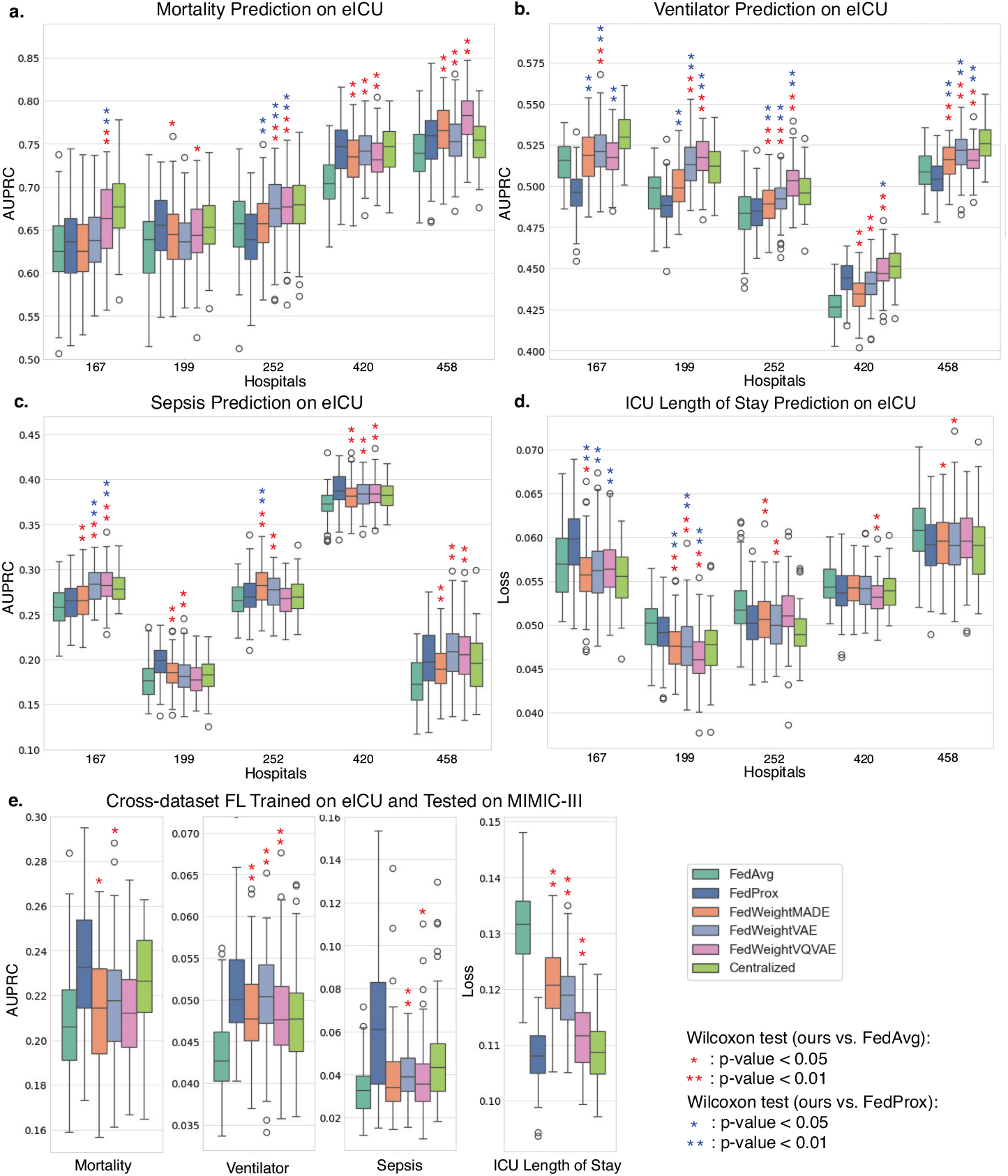
Performance comparison of federated models across clinical outcome predictions, including mortality, ventilator, sepsis, and ICU length of stay prediction. FedWeight methods were compared with FedAvg, FedProx, and a centralized model (i.e., model trained on the pooled eICU data and corrected by the covariate indicator variable for each hospital). **a** - **d**. Performance of clinical outcome predictions on eICU, which was evaluated on a bootstrap-sampled dataset from five target hospitals (167, 199, 252, 420, and 458), with mean and standard deviation computed from the bootstrap samples. **e**. Performance of cross-dataset federated model trained on eICU and evaluated on a bootstrapped test set of MIMIC-III. One-sided Wilcoxon test p-values were calculated to compare FedWeight with FedAvg and FedProx. * and ** denote p-values < 0.05 and < 0.01, respectively, for comparisons with FedAvg. * and ** indicate p-values < 0.05 and < 0.01, respectively, for comparisons with FedProx.

For mortality prediction, Hospital 458 and 420 had the best results for all methods (Figure 4a). FedWeight with the VQ-VAE density estimator provided superior results in all target hospitals (Wilcoxon test p-value < 0.05). Compared to FedProx, FedWeight performed better in Hospitals 167, 252 and 458, while FedProx had an advantage in Hospitals 199 and 420. Moreover, all FedWeight variants surpassed FedAvg in performance. They also show comparable performance to the centralized model.

For ventilator prediction, Hospital 167 and 458 demonstrate the highest AUPRC for all methods (Figure 4b), as they had the most patients and fewer imbalanced labels. Moreover, FedWeight achieved higher AUPRC scores than FedAvg, notably in Hospitals 252, 420, and 458. It also outperformed FedProx, particularly in Hospitals 167, 199, 252, and 458. Furthermore, the overall performance was close to that of the centralized model. FedWeight with the VAE density estimator demonstrated the best performance in all target hospitals.

For sepsis prediction, Hospital 420, with fewer imbalanced labels, yielded the most favourable outcomes for all methods (Figure 4c). Again, the FedWeight strategies also significantly outperformed both FedAvg and FedProx, with FedWeight using MADE and VAE providing the most accurate results in all target hospitals.

In the task of ICU length of stay prediction, most FedWeight variants yielded lower loss compared to FedAvg, demonstrated performance on par with FedProx, and closely matched the benchmark centralized model, especially in Hospital 199 (Figure 4d).

In addition, we investigated the impact of different density estimators on the final performance of FedWeight. We first observed that the sample reweights generated by the three FedWeight density estimators (MAD, VAE and VQ-VAE) exhibited high similarity, as evidenced by strong correlations among the estimated reweights across all samples (Supplementary Figure 3). Furthermore, the convergence quality of the density estimators had a noticeable effect on the performance of downstream FedWeight task models. In particular, both underfitting (due to insufficient training) and overfitting (due to excessive training) consistently led to performance degradation across various FedWeight tasks (Supplementary Figure 4).

Together, these results demonstrate that FedWeight consistently improves upon FedAvg and offers competitive or superior performance to FedProx across diverse hospitals and prediction tasks, particularly in mortality and sepsis prediction. These findings highlight the robustness and adaptability of FedWeight for real-world clinical outcome prediction in federated settings.

### Cross-dataset federated learning

In practice, each EHR dataset often requires separate access approval, making it difficult to share or centralize the data. In this scenario, models trained on each dataset can be pooled via the FL framework. To this end, we performed cross-dataset analysis by training our model on all hospitals from the eICU dataset and making predictions on MIMIC-III (Figure 4e). As expected, the models trained on eICU hospitals demonstrate worse performance on MIMIC-III (Figure 4e), compared to the performance on the held-out patients from eICU (Figure 4a-d). Even so, FedWeight outperforms FedAvg, especially in ventilator use and ICU length of stay predictions, with all FedWeight variants significantly surpassing FedAvg (Wilcoxon test p-value < 0.01) and achieving results comparable to the centralized model (i.e., model trained on the pooled eICU data and corrected by the covariate indicator variable for each hospital). Compared to FedProx, while FedWeight exhibited slightly lower average scores in mortality, sepsis, and ICU length of stay predictions, it delivered more stable performance with reduced variance. For mortality prediction, FedWeight employing MADE and VAE as density estimators achieve significantly higher AUPRC than FedAvg (Wilcoxon rank-sum test p-value *<* 0.05) (Figure 4e). In the case of sepsis prediction, FedWeight models with VAE and VQ-VAE demonstrate significantly superior performance compared to FedAvg (Wilcoxon rank-sum test p-value *<* 0.05). In addition, all FedWeight variants show statistical significance in ventilator and ICU length of stay prediction (Wilcoxon rank-sum test p-value *<* 0.01), compared to FedAvg. These findings suggest that FedWeight delivers more robust and adaptable performance, particularly in the presence of cross-dataset distribution shifts. This enhanced stability positions FedWeight as a more reliable solution for real-world federated clinical applications, where data distributions commonly vary across institutions.

### Detecting clinical features by FedWeight+SHAP analysis

We sought to assess whether FedWeight improves detecting relevant features for the clinical outcome predictions using Shapley Additive Explanations (SHAP) [49]. As a reference, we computed the SHAP values of the centralized model. We used the Pearson correlation of the SHAP values between the federated model and the centralized models as the evaluation metric. We first performed the experiments using hospitals within the eICU dataset. FedWeight demonstrated a superior correlation of SHAP values compared to FedAvg across almost all hospitals, especially in predicting ventilator use. Specifically, for mortality prediction, all the FedWeight methods significantly outperform the baseline method (Wilcoxon test p-value < 0.01), except for Hospital 458 (Figure 5a). Moreover, the FedWeight model using MADE and VAE density estimator demonstrated significantly higher correlation of SHAP values in all hospitals (Wilcoxon test p-value < 0.01). For ventilator prediction, FedWeight using VAE and VQ-VAE density estimator significantly outperformed FedAvg in all target hospitals (Wilcoxon test p-value < 0.01) (Figure 5b). Regarding sepsis prediction, FedWeightVAE demonstrated the most significant results in all target hospitals (Wilcoxon test p-value < 0.05) (Figure 5c). For the length of stay prediction, FedWeightMADE demonstrated significantly stronger correlations with the benchmark across all target hospitals, whilst FedWeightVAE showed significantly higher correlations in Hospital 167, 252, 420, 458 (Wilcoxon test p-value < 0.01) (Figure 4d). To conclude, our experiments on the eICU dataset demonstrated that FedWeight SHAP values are more consistent with the benchmark SHAP values. As SHAP values quantify feature importance, this suggests that FedWeight is more effective in capturing influential features.

**Figure 5:**
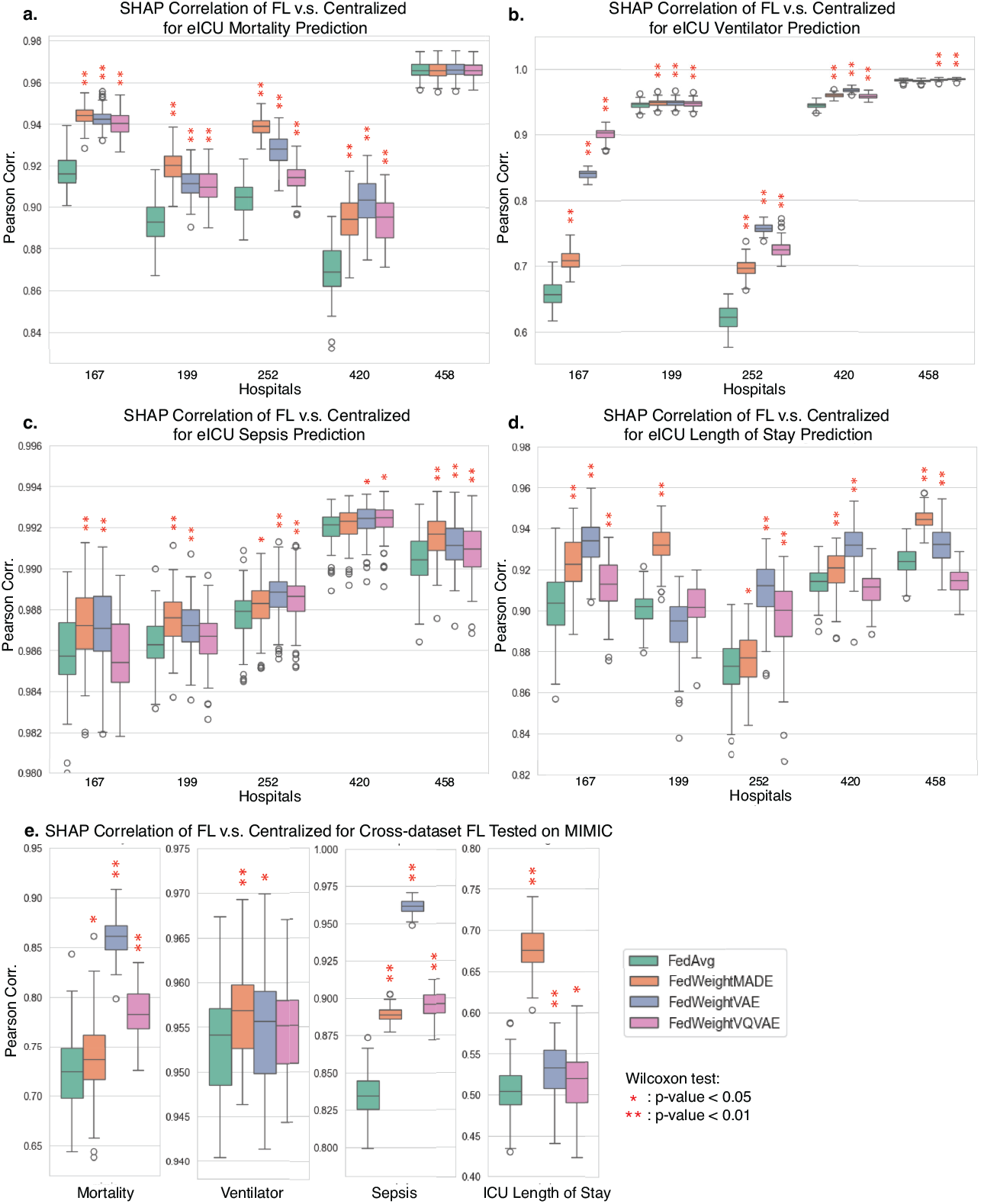
Comparison of FedWeight and FedAvg in capturing influential features. SHAP values were leveraged to identify feature importance, calculated from test data fed into the trained model. For mortality and ICU length of stay prediction, SHAP values were summed across all samples. For ventilator and sepsis prediction, SHAP values were aggregated across all time windows and samples, resulting in a one-dimensional vector of feature importance. We then calculated the Pearson correlation of feature importance between the federated and centralized model. **a** - **d**. Pearson correlation of SHAP-based feature importance for clinical outcome predictions in eICU. The models were validated on the bootstrapped test sets of five target hospitals (167, 199, 252, 420, 458). **e**. Pearson correlation of SHAP-based feature importance for cross-dataset federated learning, where models were trained on eICU, and the correlation was computed on the bootstrapped test set of MIMIC-III. One-sided Wilcoxon test p-values were calculated against the baseline. * denotes the p-values *<* 0.05, and ** represents the p-values *<* 0.01.

We further performed a cross-dataset analysis, using eICU hospitals as the source and MIMIC-III as the target. FedWeight demonstrated significantly higher correlations in predicting mortality, sepsis, and ICU length of stay (Wilcoxon test p-value *<* 0.01) (Figure 5e). Notably, FedWeight utilizing the VAE as the density estimator exhibited the best performance in mortality and sepsis prediction, underscoring its capacity to capture features highly associated with a patients’ length of stay in the ICU.

### Top drugs and lab tests attributed to the clinical outcomes

Given the strong quantitative results for the feature attributions, we turn to individual drugs and lab tests that exhibit high SHAP values based on the best FedWeight model, namely FedWeightVAE.

#### Mortality

We leveraged drug administration and lab tests from the initial 48 hours of ICU admission to predict patient mortality beyond this period. The drugs of the highest correlation with patient mortality are predominantly vasopressors or anaesthetics (Figure 6a). Specifically, glycopyrrolate is the foremost drug in mortality prediction, which reflects its role as an anticholinergic agent to manage respiratory secretions and mitigate vagal reflexes in critically ill patients during surgery [50]. Its association with mortality may serve as a proxy marker for high-acuity clinical scenarios, highlighting its relevance as a potential marker of severe physiological compromise in ICU settings. Following glycopyrrolate, vasopressors like vasopressin epinephrine, and phenylephrine emerge as critical prior to patients mortality, highlighting their role as primary stress hormones typically administered in the context of life-threatening conditions such as septic shock, cardiogenic shock, or profound hypotension [51–54]. Therefore, this correlation suggests that the necessity for vasopressor support often signals an advanced stage of critical illness, marked by high acuity and poor prognostic outcomes. After vasopressors, we also observed the administration of morphine prior to patient mortality in the ICU, which is likely attributable to its role in palliative care and the management of refractory pain and dyspnea in critically ill patients [55].

**Figure 6:**
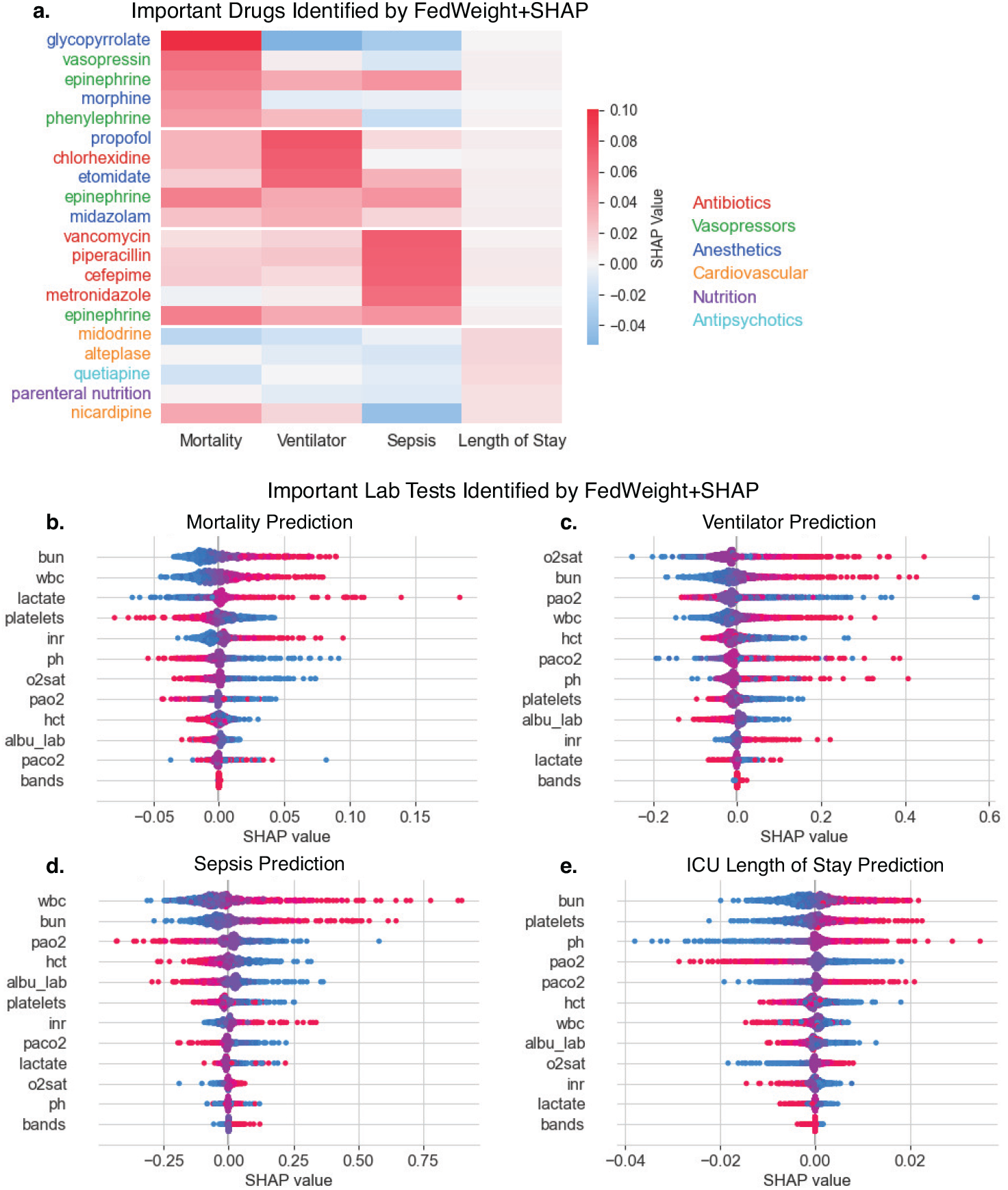
Feature importance of drug and lab tests for clinical outcome predictions identified by FedWeight+SHAP. **a**. SHAP values of the top 5 most important drugs identified by FedWeight for each clinical outcome. We computed SHAP values for each drugs present on target hospital. For mortality and ICU length of stay prediction, SHAP values were summed across all patients, while for ventilator and sepsis prediction, they were summed across both time windows and samples, resulting in a one-dimensional feature importance vector. For each clinical outcome, the top 5 most important drugs were selected, visualized with colour intensities indicating their feature importance. **b** - **e**. SHAP values of the top 12 most important lab tests for each clinical outcome, identified by FedWeightVAE. Each dot represents a sample from the target hospital. Features are ordered by their mean absolute SHAP value.

We identified a significant correlation between elevated blood urea nitrogen (BUN) levels and subsequent patient mortality, highlighting its utility as a pivotal biomarker for mortality risk in critical care (Figure 6b). Elevated BUN reflects underlying pathophysiological processes, including renal insufficiency, systemic hypoperfusion, and heightened protein catabolism, which are key indicators of severe illness [56–60]. Therefore, this association underscores the prognostic significance of BUN in stratifying patient risk and guiding therapeutic interventions. Additionally, increased lactate levels are another profound indicators of mortality. This verifies the existing medical knowledge, as elevated lactate is a key clinical criterion for tissue hypoxia in critically ill patients, which is strongly associated with ICU mortality [61–64].

#### Ventilator use

Since most patients initiate ventilation within 72 hours of ICU admission, we established a 72-hour observation window, further segmented into six 12-hour intervals. We aim to utilise drug administration and lab test results from each interval to predict ventilator use in the next. Additionally, we observed that 65.5% of patients in the eICU dataset experience multiple episodes of mechanical ventilation during their ICU stay. As a result, ventilator treatment may appear in multiple intervals for a single patient. Among all the drugs, propofol stands out for ventilator prediction, which is commonly administered in critical care to facilitate patient-ventilator synchrony, minimize agitation, and ensure tolerance of invasive respiratory support [65, 66]. However, since propofol is commonly administered during intubation, its association with subsequent ventilator use likely stems from multiple ventilation episodes throughout the ICU stay. Following propofol, our model identified chlorhexidine as a drug administered to ICU patients who subsequently require mechanical ventilation, which facilitates the identification of patients at high risk for ventilation, while enabling medical practitioners to proactively prepare for ventilatory support. This identification verifies the established clinical knowledge. As a broad-spectrum antiseptic with bactericidal properties, chlorhexidine is widely utilized in oral care protocols within the ICU to mitigate the microbial colonization of the oropharynx and trachea, which are primary precursors to ventilator-associated pneumonia (VAP) [67–69]. Our model also identified that etomidate is the third most commonly administered drug prior to ventilator use. Specifically, etomidate is a rapid-onset, short-acting intravenous anaesthetic commonly employed before endotracheal intubation. Although its administration might be associated with adrenal suppression, etomidate is particularly suitable for hemodynamically unstable patients due to its minimal impact on blood pressure [70, 71].

For ventilator prediction, elevated arterial blood gas (ABG) parameters, including oxygen saturation (O2sat), demonstrate a strong correlation with the subsequent initiation of ventilatory support (Figure 6c). While this may appear counterintuitive, as oxygen saturation measurements are recorded before the initiation of ventilation, one would anticipate these parameters to be elevated only after ventilator treatment begins. Multiple episodes of mechanical ventilation could explain this phenomenon. This pattern could also arise from the initial use of high-flow oxygen or non-invasive ventilation (e.g., CPAP or BiPAP) as a preparatory measure before transitioning to invasive mechanical ventilation. Therefore, it is conceivable that patients undergo an initial period of ventilatory support, exhibit elevated oxygen saturation values, and subsequently require re-initiation of mechanical ventilation. Moreover, we also observed the elevated blood urea nitrogen (BUN) levels are highly correlated with the subsequent ventilatory support. Such elevations often stem from renal insufficiency, hypovolemia, or heightened protein catabolism associated with conditions like sepsis or multiorgan dysfunction. These pathophysiological states frequently precede respiratory failure, necessitating the commencement of mechanical ventilation. Therefore, markedly elevated BUN levels may serve as an ICU risk marker which often guides decisions on fluid management, dialysis, and ventilation timing.

#### Sepsis

Similar to ventilation use, we designed a 72-hour observation window comprising six 12-hour intervals. We aim to employ drug administration and lab tests from each interval to predict sepsis diagnosis in the subsequent interval. Confirming the existing medical knowledge and literature, antibiotics such as vancomycin, exhibit the highest overall feature attribution [72, 73], followed by piperacillin [74], cefepime [75], and metronidazole [76]. Notably, our model make prediction based on drugs administered prior to the sepsis diagnosis. Given that approximately 90% of sepsis cases are community-onset [77], with most patients presenting infection symptoms prior to sepsis diagnosis, it is routine for patients to receive these antibiotics before a formal diagnosis of sepsis.

For sepsis prediction, the white blood cell (WBC) count holds the highest SHAP value (Figure 6d), underscoring its strong association with septicemia and septic shock. Leukocytosis generally reflects the immune system’s activation in response to bacterial infection, driven by pro-inflammatory cytokines [78]. Therefore, clinicians may leverage WBC with other laboratory markers, including creatinine, platelet count, and lactate, for early diagnosis and monitoring of the condition.

#### ICU length of stay

Using drug administration and lab tests from the first 48 hours of ICU admission, we aim to predict the remaining ICU stay duration. Patients receiving cardiovascular drugs, including midodrine, alteplase, and nicardipine, tend to have prolonged ICU stays (Figure 6a). This indicates that patients with cardiac surgery often require prolonged monitoring and extended care, as supported by existing research [79].

We observed an elevated platelet count as a strong indicator of prolonged ICU length of stay (Figure 6e). Specifically, thrombocytosis serves as a surrogate marker of heightened inflammatory activity and immune system activation. This hyperactive platelet response may signify the severity of the underlying pathology, such as infection, malignancy, or surgical recovery, all of which demand intensive and sustained care. Additionally, elevated platelet levels are indicative of a hypercoagulable state, predisposing patients to thrombotic complications, such as deep vein thrombosis or pulmonary embolism, which necessitates prolonged ICU stay for vigilant monitoring and therapeutic intervention [80]. Therefore, the association of thrombocytosis with prolonged ICU duration underscores its role as a biomarker of systemic stress and disease severity, providing insights into patient prognosis and resource allocation in the ICU.

Together, our combined FedWeight+SHAP analysis detected known drugs and lab tests associated with the clinical outcome predictions. This identification of influential variables warrants future investigation to explore causal mechanisms for the nuanced clinical associations identified in this analysis.

### Mortality-associated latent topics captured by FedWeight-ETM

In the aforementioned experiments, FedWeight demonstrated exceptional performance in supervised prediction tasks. To identify sets of correlated clinical features in an unsupervised setting, topic models [81, 82] are natural choices as they can capture underlying patterns in high-dimensional EHR data [83, 84]. To achieve this, we incorporated FedWeight into Embedded Topic Model (ETM) [82] (see “Federated embedded topic model” in **Methods**), leveraging its ability to learn a low-dimensional semantic representation of clinical features while preserving patient privacy in a FL setting. First, we developed FedAvg-ETM, which shares the encoder weights and clinical feature embeddings between silos (i.e., eICU and MIMIC-III) to infer robust latent topics from distributed EHR data (Figure 7a). To address covariate shifts, we further extended FedAvg-ETM to FedWeight-ETM (Figure 7b). Quantitatively, we benchmarked the performances of non-federated ETM, FedAvg-ETM, and FedWeight-ETM by predicting readmission mortality using the corresponding topic mixtures as inputs to a logistic regression classifier. Overall, FedAvg-ETM and FedWeight-ETM demonstrated comparable performance and outperformed the baseline non-federated ETM across different topic numbers (Supplementary Figure 5).

**Figure 7:**
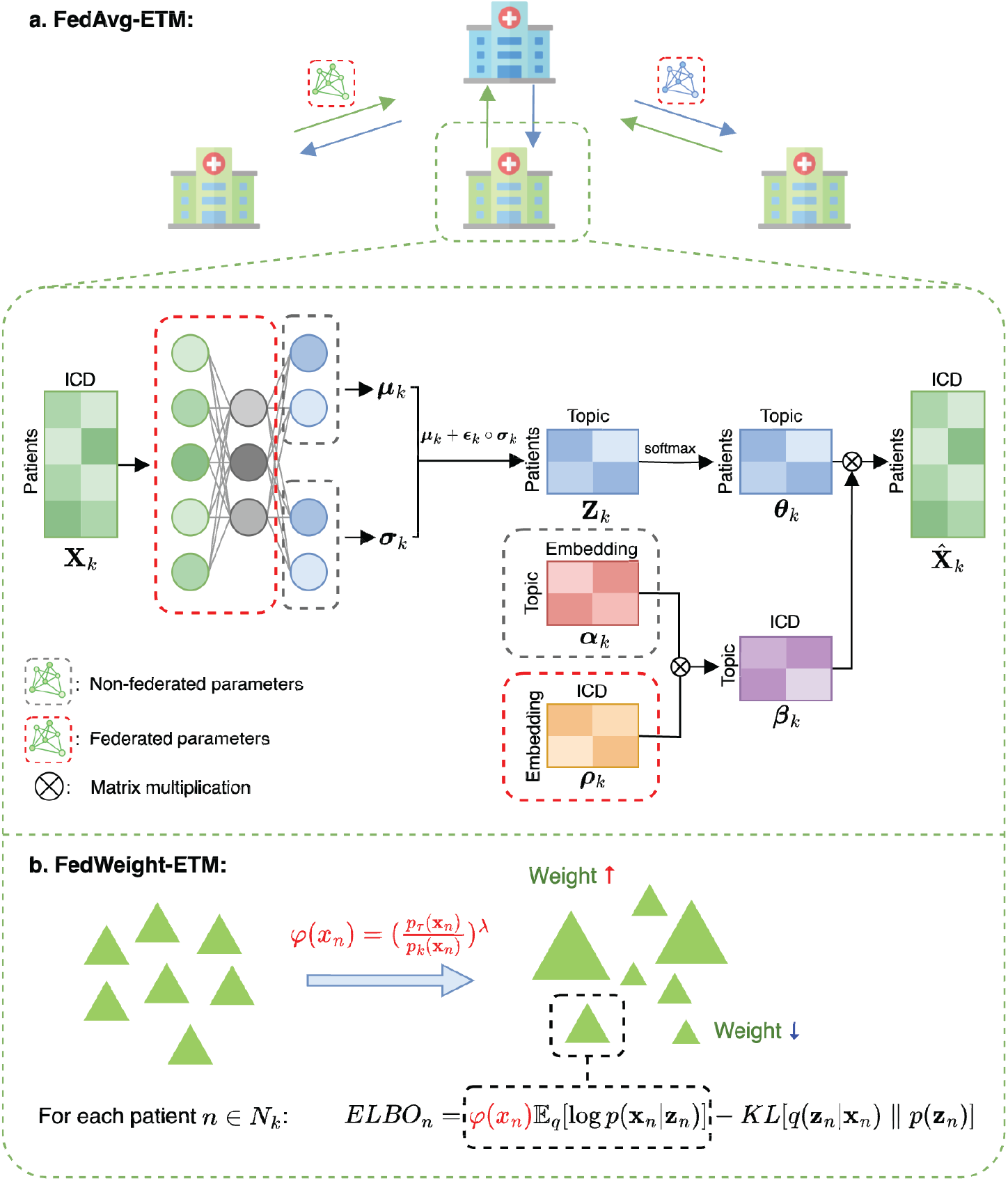
Federated latent topic modelling. **a**. FedAvg-ETM. In each local hospital *k* ∈ *K*, **X**_*k*_ is input into the encoder, whose output goes into two separate linear layers and produces ***µ***_*k*_ and ***σ***_*k*_. Then, through the re-parameterization trick, we obtain the latent representation **Z**_*k*_. After applying the softmax function, **Z**_*k*_ gives the patient-topic distribution ***θ***_*k*_. The learnable topic embedding ***α***_*k*_ and ICD embedding ***ρ***_*k*_ generate the topic-ICD mixture ***β***_*k*_. Then, ***β***_*k*_ is multiplied with ***θ***_*k*_ to reconstruct the input. During federated averaging, only the encoder network and the ICD embedding are uploaded to the target hospital for aggregation, whilst all other model parameters are kept locally updated. **b**. FedWeight-ETM. FedWeight-ETM builds on FedAvg-ETM by applying a re-weight to the log-likelihood term of the ELBO function for each patient.

We evaluated FedWeight-ETM’s ability to capture topics related to patient mortality. We identified the top 5 topics that are significantly associated with patient mortality. Figure 8a and b display the top three-digit International Classification of Diseases (9th revision)(ICD) diagnostic codes [85] under each topic supported by the Fisher-exact test p-values for eICU and MIMIC-III data, respectively. For eICU, Topic 16 is the most significant topic with the top ICD code being chronic renal failure (ICD-585). Indeed, renal failure disrupts fluid, electrolyte, and metabolic equilibrium and ultimately systemic homeostasis. For critically ill patients, it exacerbates comorbidities such as sepsis and is a hallmark of multiorgan dysfunction syndrome (MODS), perpetuating systemic inflammation and hemodynamic instability, which increases the risks of patient mortality [86, 87]. As the second significant topic, Topic 31 involves cardiac dysrhythmias (ICD-427), which has direct impact on hemodynamic stability and causes critical conditions such as myocardial ischemia, heart failure, or sepsis. Moreover, ventricular arrhythmias and atrial fibrillation can lead to reduced cardiac output, tissue hypoperfusion, and multiorgan dysfunction in critically ill patients [88]. As the third focus, Topic 28 involves acute renal failure (ICD-584). We observed that while acute renal failure is strongly associated with mortality, its correlation is weaker than that of chronic renal failure. This may be due to its potential reversibility with timely medical intervention [89].

**Figure 8:**
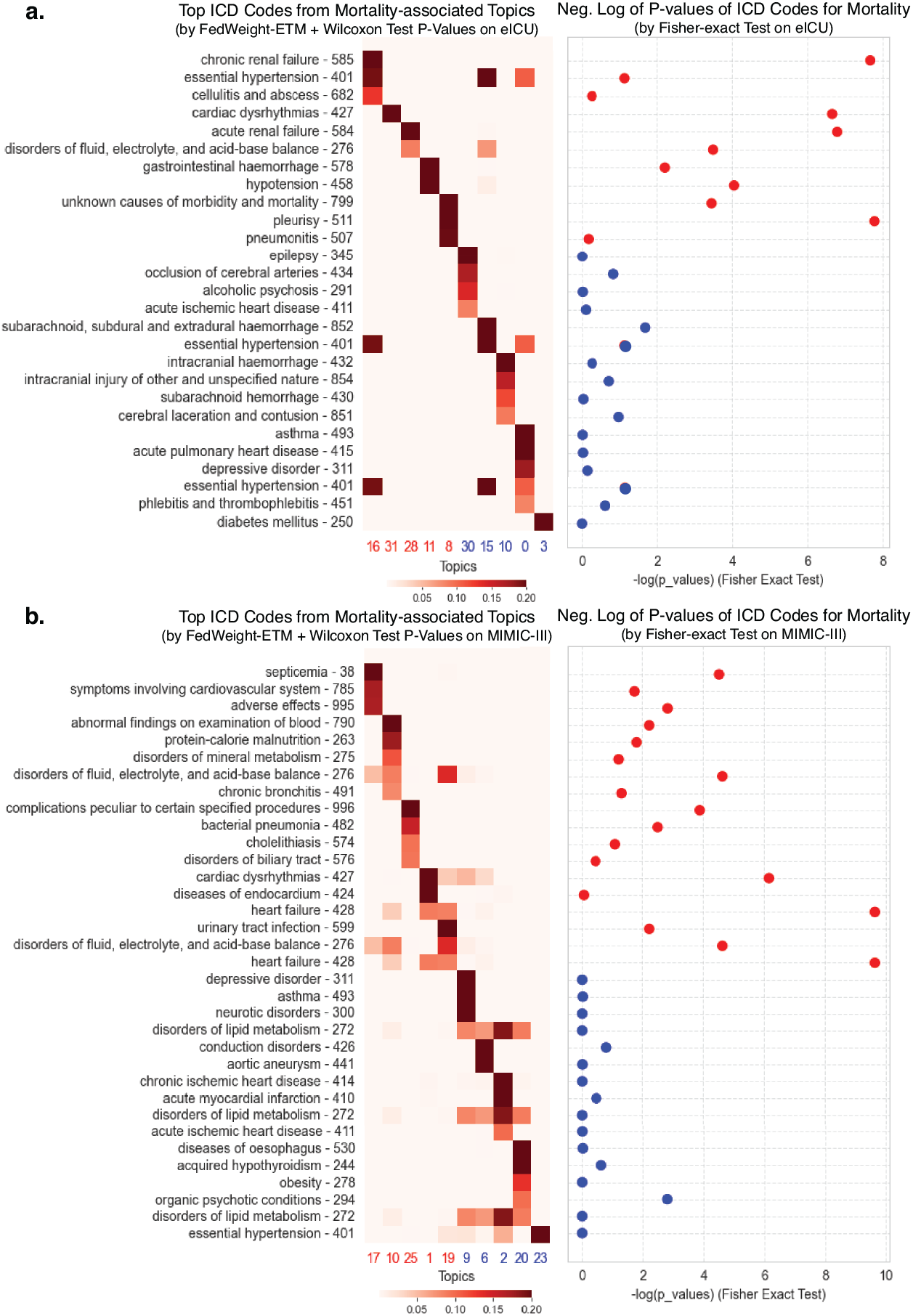
Relationship between the most and least mortality-associated topics and ICD codes identified by FedWeight-ETM. We identified the top 5 mortality-associated topics based on Wilcoxon test on topic proportions between deceased and surviving patients. The colour intensity of the heatmap indicates the probability of an ICD codes within a given topic. We selected the top ICD codes whose probability is greater than 0.08. These ICD codes were validated by Fisher’s exact test p-values on the eICU and MIMIC-III data, visualized in a scatter plot for the most and least mortality-associated ICD codes. **a**. Mortality-associated topics and ICD codes on eICU. **b**. Mortality-associated topics and ICD codes on MIMIC-III.

We also observed clinically meaningful topics inferred from the MIMIC-III dataset (Figure 8b). Namely, Topic 17 is characterized by septicemia (ICD-38), which triggers widespread endothelial damage, capillary leakage, and coagulopathy, culminating in septic shock and multi-organ failure [90]. Furthermore, for Topic 10, the top disease identified by our model is abnormal findings on examination of blood (ICD-790). Severe abnormalities in blood parameters—such as metabolic acidosis, electrolyte imbalances, coagulation abnormalities, and hematologic dyscrasias—can precipitate multi-organ failure, hemodynamic instability, and increased susceptibility to infections [91]. These abnormalities often reflect underlying pathophysiological insults, such as sepsis, acute kidney injury, or haematological malignancies, which all increase the risks of patient mortality. As the third significant topic, Topic 25 is identified as complications peculiar to certain specified procedures (ICD-996). These complications involve device-related infections, graft failures, prosthetic malfunctions, and post-surgical hemorrhage, which can precipitate systemic instability, multi-organ failure, or sepsis. This indicates that FedWeight-ETM is able to efficaciously capture semantically meaningful topics, thus helping to uncovering clinically relevant patterns in healthcare data. The association of these medical conditions with mortality underscores the necessity of rigorous monitoring, prompt identification, and timely intervention to mitigate these risks and improve patient survival.

## Discussion

FL is a promising approach for leveraging decentralized EHR data. However, FL notoriously suffers from the covariate shift issue, where data distributions differ significantly across clinical sites. These differences in demographics, clinical practices, and data collection processes may lead to significant performance degradation of the shared model when deployed for a target population. To mitigate this issue, we propose FedWeight, where we probabilistically re-weight the patients from the source hospitals. Intuitively, samples more similar to the target distribution receive higher weights, thus contributing more during training, whereas those less similar are assigned lower weights, thereby contributing less during training. This approach ensures that the data more relevant to the target distribution has a more significant impact on model training, thus aligning the trained model with the data distribution of the target hospital and effectively addressing covariate shifts in FL environments.

We conducted extensive experiments by FL across hospitals within the eICU dataset and between the eICU and MIMIC-III datasets. Our approach demonstrates the following strengths: (1) enhances the generalization of FL classifiers for predicting clinical outcomes such as mortality, sepsis, ventilator usage, and ICU length of stay; (2) uncovers subtle yet significant drugs and lab tests associated with clinical outcomes; and (3) identifies relationships between diseases, involving renal and heart failure, and future mortality at ICU readmission. Compared to FedAvg and FedProx, FedWeight achieved more stable performance with lower variance across runs, even though FedProx occasionally attained higher average scores in specific tasks. This robustness under covariate shift highlights FedWeight’s suitability for real-world federated clinical applications, where differences in patient demographics and treatment practices are common. Moreover, FedWeight maintains a lightweight design with minimal computational overhead relative to FedAvg, making it practical for deployment at scale. We have also theoretically analyzed its convergence properties (refer to Supplementary Note 1) and empirically validated its training stability. These findings underscore the scalability and reliability of FedWeight when applied to decentralized clinical data affected by distributional shift.

In future work, we aim to theoretically investigate why certain density estimators (e.g. VAE) perform better in specific prediction tasks. We also plan to develop density estimators and federated models that perform well even on small datasets. Furthermore, we now aggregated patient clinical data across all time points to estimate density. We will also explore density estimators designed for time series data. Moreover, although our method enhances performance on the target hospital, its ability to generalize to entirely new or highly heterogeneous sites remains uncertain. A potential future direction could involve systematically analyzing the degree of data heterogeneity under which FedWeight outperforms other methods. We will establish quantitative benchmarks to rigorously assess its effectiveness across different levels of distribution shifts. Besides, since some patients experience multiple ventilation episodes, medications administered during treatment may be mistakenly identified as predictors of future treatments. We aim to mitigate this spurious correlation in our future work. Moreover, to further protect patients’ privacy, we aim to incorporate additional privacy-preserving techniques, such as differential privacy, into our future research directions. Finally, we will integrate our method into existing open-source Software Development Kit (SDK) such as Flower [92], Federated AI Technology Enabler (FATE) [93], FedScale [94], as well as NVIDIA Federated Learning Application Runtime Environment (NVIDIA FLARE) [95], for practical application in real-world health institutions.

## Methods

### FedWeight

#### Problem formulation

Assuming there are *K* source hospitals and one target hospital *τ* in the federated

network. For each source hospital *k* ∈ *K*, we have input data 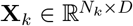 and labels 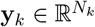, where *N*_*k*_ is the number of patients in hospital *k* and *D* is the feature size, and 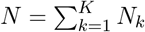 is the total number of patients in the federated network. The notations and their corresponding descriptions are outlined in Supplementary Table 3.

#### Federated Learning

Before delving into FedWeight, we first described the baseline Federated Average (FedAvg) [3], whose training process is as follows:

- Local computation: Each source hospital *k* ∈ *K* updates its model parameters **w**_*k*_ on its local data.
- Parameter sharing: Then, the source hospital *k* sends its updated parameters **w**_*k*_ to the target hospital *τ*.
- Aggregation at the target hospital: The target hospital *τ* receives these updated parameters and aggregates them to update the global model, which is then sent back to the source hospitals for the next round training.

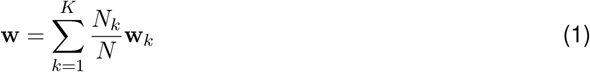

#### Weighted log-likelihood

However, FedAvg does not account for the covariate shift issue. Specifically, covariate shift can happen when the source and target hospitals may have different local resources / clinical practices: *p*_*k*_(**X**) ≠ *p*_*τ*_ (**X**) while clinical outcomes given certain drugs remain similar across hospitals: *p*_*k*_(**y**|**X**) = *p*_*τ*_ (**y**|**X**). To mitigate this issue, we may employ the weighted log-likelihood algorithm [13], which modifies the standard log-likelihood calculation by assigning weights to different data points. Samples more similar to the target distribution are given higher weights, thus contributing more during training, while those less similar are assigned lower weights, thereby contributing less during training. Consequently, this approach ensures that the trained model gives preference to data which are more relevant to the target distribution, thus addressing covariate shift in FL environments.

The weighted log-likelihood for hospital *k* can be expressed mathematically as follows:

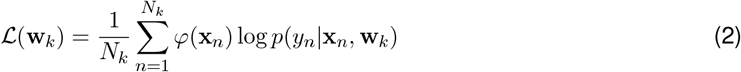

where 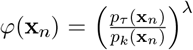 denotes the weight assigned to the *n*-th data point, *p*_*τ*_ represents the density estimator trained on target hospital *τ*, and *p*_*k*_ denotes the density estimator trained on source hospital *k*. Furthermore, *λ* is a hyper-parameter that controls the degree of re-weighting. Moreover, we employed various density estimators such as MADE, VAE, and VQ-VAE on account of their suitability in estimating the underlying probability distributions.

We developed FedWeight by incorporating the weighted log-likelihood algorithm into FL settings. Therefore, the log-likelihood for the entire federated network can be expressed as follows:

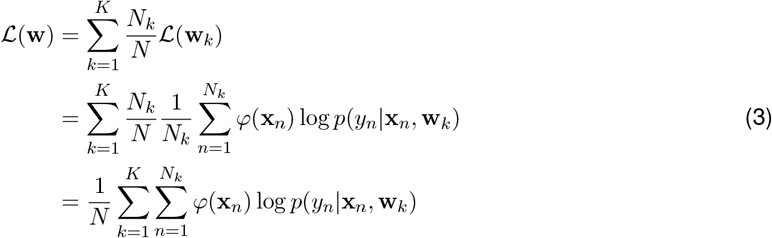

#### Density estimation

To calculate the re-weight *φ*(**x**_*n*_), we need to effectively train the density estimator *p*_*k*_ and *p*_*τ*_, which is described below.

##### (1) Masked Autoencoder for Density Estimation (MADE)

The Masked Autoencoder for Density Estimation (MADE) is a neural network model for efficient density estimation in high-dimensional data [40]. It modifies the traditional autoencoder architecture by applying masks to its connections, ensuring that the output at each unit satisfies the autoregressive property, thus effectively modelling the input data’s joint distribution.

The density estimation using the autoregressive property is calculated as follows:

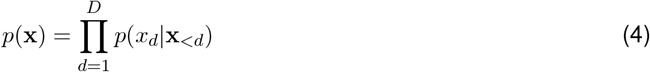

To achieve the autoregressive property, the conventional autoencoder architecture is modified by masking some of the model weights. Therefore, the MADE training algorithm consists of the following three steps:

- Number Assignment: Initially, a unique integer ranging from 1 to *D* is sequentially assigned to each input and output unit of the autoencoder model. Moreover, every hidden unit is randomly allocated an integer within the range of 1 to *D* − 1, inclusive.
- Mask Construction: Assuming the autoencoder model consists of *L* layers, let 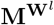 represent the mask matrix between layer *l* and its preceding layer *l* − 1, except for the output layer. We connect the unit *k*^′^ in layer *l* and the unit *k* in its preceding layer *l* − 1 only if the assigned integer *m*^*l*^(*k*^′^) is greater than or equal to *m*^*l*−1^(*k*); in all other cases, we apply masks. Therefore, this mask matrix is calculated as follows:

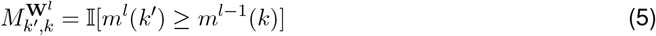 Let **M**^**V**^ denote the mask matrix between the output layer *L* and the last hidden layer *L*−1. Specifically, we connect the output unit *d*^′^ and the unit *d* in the last hidden layer only if the assigned integer *m*^*L*^(*d*^′^) is **strictly** greater than *m*^*L*−1^(*d*); in all other cases, we apply masks. Therefore, this mask matrix is calculated as follows:

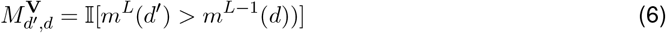
- Output Calculation: Then the autoencoder output 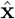 is computed as follows:

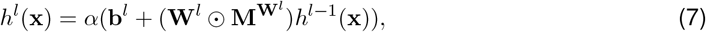

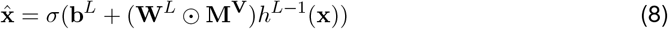

where the masks 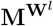 and **M**^**V**^ are applied through element-wise multiplication with their respective model weights **W**^*l*^ and **W**^*L*^. **b**^*l*^ and **b**^*L*^ are the bias for the model hidden and output layers. *α* is the activation function in the hidden layer, while *σ* denotes the sigmoid function.

Given the output 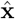, we find the model parameters to maximize the reconstruction likelihood.

### (2) Variational Autoencoder (VAE)

Variational Autoencoder (VAE) is a generative machine learning model that uses deep neural networks to encode data into a latent space and then decode it back, enabling tasks like data generation and density estimation [41]. Specifically, a VAE model consists of two coupled components: an encoder model *q*_*φ*_(**z**|**x**), and a decoder model *p*_*θ*_(**x**|**z**). During training, the encoder maps the observations **x** to an approximate posterior over the latent variables, parameterized by a mean ***µ*** and variance ***σ***^2^. Instead of directly sampling the latent representation **z** from the encoded latent distribution *q*_*φ*_(**z**|**x**), which is non-differentiable, **z** can be expressed as **z** = ***µ*** + ***σ*** ⊙ ***ϵ***, where ***ϵ*** ~*𝒩* (0, **I**). Finally, the decoder ingests the latent representations **z** to reconstruct the initial data 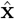.

Furthermore, the decoder parameters *θ* and the variational parameters *φ* are learned by maximizing the Evidence Lower Bound (ELBO), where *p*(**z**) denotes a standard normal distribution.

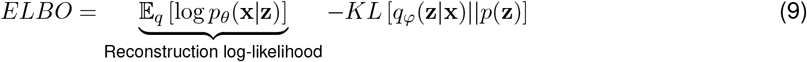

Based on log *p*(**x**) = *KL*[*q*_*φ*_(**z**|**x**)||*p*(**z**|**x**)] + *ELBO*, when the model is converged, we expect the variational distribution *q*_*φ*_(**z**|**x**) approaches the true posterior distribution *p*(**z**|**x**), leading to their KL divergence converges to zero. Consequently, we may leverage ELBO in density estimation, as log *p*(**x**) approaches to ELBO when the model is converged.

### (3) Vector Quantized Variational Autoencoder (VQ-VAE)

In addition to the aforementioned density estimators, we may also utilize the Vector Quantized Variational Autoencoder (VQ-VAE) to estimate density [42]. Compared with traditional VAE that utilizes continuous latent representations, VQ-VAE employs a discretized latent space. Moreover, VQ-VAE uses categorical distributions for both posteriors *q*(**z**|**x**) and priors *p*(**z**).

Similar to the VAE architecture, VQ-VAE also contains an encoder and decoder. Further, VQ-VAE maintains a codebook **E** = {**e**_1_, **e**_2_, …, **e**_*C*_}, where *C* is the total codewords. For each codeword **e**_*c*_ ∈ ℝ ^*H*^,*H* denotes the codeword dimension.

When training VQ-VAE, the encoder takes **x** as input and generates **z**_*e*_. Then **z**_*e*_ goes into the codebook **E** to find the index of the nearest codeword *ĉ* = argmin_*c*∈*C*_ ||**z**_*e*_ − **e**_*c*_||^2^. Then we use this index to construct the one-hot encoded latent representation **z** = [*z*_*c*_]_×*C*_, where *z*_*c*_ = 𝕀 [*c* = *ĉ*]. Subsequently, the decoder takes **e**_*ĉ*_ as input and reconstructs the original input data 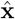. Finally, we may leverage ELBO in density estimation.

#### FedWeight algorithm design

The FedWeight training process is as follows:

- Density estimator training: At the beginning of the training process, the target hospital *τ* trains a density estimator *p*_*τ*_. Meanwhile, each source hospital *k* ∈ *K* also trains its own density estimator *p*_*k*_. We selected specific density estimator models, including MADE, VAE, and VQ-VAE on account of their suitability in estimating the underlying probability distributions (Figure 3a).
- Sharing of target density estimator: Then the target hospital distributes its density estimator *p*_*τ*_ to all the source hospitals (Figure 3b).
- Re-weighted local model training: Upon receipt *p*_*τ*_, each source hospital *k* ∈ *K* computes the reweight 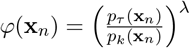 for patient *n* ∈ *N*_*k*_, where *λ* is a hyper-parameter that controls the degree of re-weighting. Leveraging such re-weight, the source hospital *k* trains its own model locally. Then, the parameters of such model **w**_*k*_ is further sent to the target hospital *τ* (Figure 3c).
- Aggregation at the target hospital: The target hospital *τ* receives these updated parameters and aggregates them to update the global model, which has better generalization capabilities when applied to the target hospital’s data (Figure 3d).

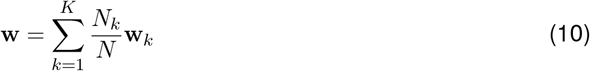

FedWeight effectively adapts the model trained on source hospital data to a target hospital. However, there may be scenarios in which hospitals must mutually benefit from each other’s data distributions. To address the collaboration among these hospitals, we developed a Symmetric FedWeight training paradigm. For simplicity, we assumed a federated network with two hospitals, namely A and B. During local training at Hospital A, Hospital B is treated as the target to estimate the reweighting ratios. Conversely, when training at Hospital B, Hospital A serves as the target for reweighting. This symmetric approach allows both hospitals to benefit from each other by adapting to their respective data distributions (Figure 3e-h).

FedWeight employs task-specific models, including LSTM for sequence prediction and VAE for density estimation. Detailed model architectures are provided in Supplementary Note 2. Moreover, to protect patients’ privacy, only the parameters of the models are exchanged within the federated networks, ensuring no transmission of raw data.

We also analyzed FedWeight’s convergence behaviour under covariate shifts and found that it achieves a convergence rate of *O*(1*/T*), aligning with FedAvg. The detailed convergence analysis is provided in Supplementary Note 1.

### Federated embedded topic model

*ETM*. ETM is a generative model of documents that learns interpretable topics and word embeddings and is robust to large vocabularies. For each patient diagnosis data point **x**_*n*_, the encoder, parameterized by **w**_*θ*_, maps it to ***µ***_*n*_ and log 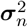 through two distinct linear layers, parameterized by **w**_*µ*_ and **w**_*σ*_.

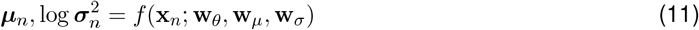

Through the re-parameterization trick, we obtain the latent represatation **z**_*n*_, which eventually outputs the patient-topic mixture ***θ***_*n*_ via a softmax operation across all latent topics.

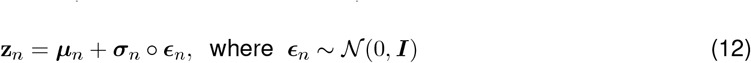

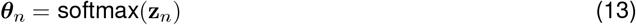

The decoding process involves the multiplication of two learnable matrices: the topic embedding matrix ***α*** and the ICD embedding matrix ***ρ***. The operation yields a topic-ICD mixture ***β***, which represents the probabilistic association between topics and ICD codes. Eventually, ***β*** is multiplied by ***θ***_*n*_, resulting in the reconstruction of the input 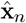.

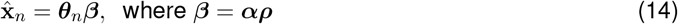

ETM is trained by maximizing the following ELBO function:

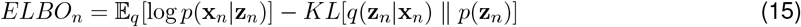

#### FedAvg-ETM

The topics are not identifiable between hospitals, preventing directly model averaging. Instead, we aggregate only **w**_*θ*_ and ***ρ***, while **w**_*µ*_, **w**_*σ*_, and ***α*** are kept locally updated (Figure 7a). Specifically, the FedAvg-ETM training process is as follows:

- Local computation: Each source hospital *k* ∈ *K* updates its local model parameters

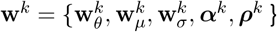
- Parameter sharing: Then, the source hospital *k* sends the non-topic-associated model parameters 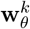 and ***ρ***^*k*^ to the target hospital *τ*.
- Aggregation at the target hospital: The target hospital *τ* receives these updated parameters and aggregates them to update the global model, which is then sent back to the source hospitals for the next round of training.

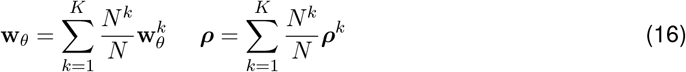

#### FedWeight-ETM

Variations in clinical practices, patient demographics, and data collection methods, may engender covariate shifts in clinical data, which hinders FedAvg-ETM from effectively uncovering semantically meaningful latent topics. To address this challenge, we proposed FedWeight-ETM, which integrates the FedWeight framework with the ETM model. Specifically, we assign the reweighting ratio 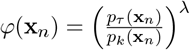 to the likelihood term of the ELBO function of the *n*-th data point (Figure 7b), leading to the weighted ELBO function:

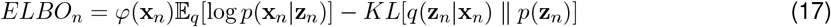

### Data preprocessing

We applied our method to the eICU Collaborative Research Database with data from 208 hospitals and 200,859 patients, and the MIMIC-III dataset. For the eICU dataset, we selected 10 hospitals with the most patients: Hospitals 167, 420, 199, 458, 252, 165, 148, 281, 449, and 283. The dataset includes 1,399 distinct drugs, which were binarized for model analyses; a value of “1” indicates the patient received the drug, whereas “0” signifies no administration. Since patients typically receive only 1% to 2% of the total drug catalogue, this binarization results in a sparse matrix with a predominance of “0” values in the dataset. In addition to drug data, our model incorporates patient demographics (age, sex, BMI, and ethnicity) to enhance predictive accuracy. To avoid null or incomplete demographic records, we excluded samples with missing values; however, the proportion of such records is very low compared to the full dataset (Supplementary Figure 6). Age was encoded into eight categories: <30, 30-39, 40-49, 50-59, 60-69, 70-79, 80-89, and >89 years. Sex was dichotomized, with “1” representing males and “0” indicating females. BMI was categorized into four groups: underweight (<18.5), normal (18.5-24.9), overweight (25-29.9), and obesity (≥ 30) [96]. Ethnicity was classified into five categories: Caucasian, African American, Hispanic, Asian, and Native American, reflecting the diversity within the patient population. For MIMIC-III dataset, we removed the newborn patients. Moreover, given the small sample size, we excluded the MIMIC patients in Cluster 2 for simplicity (Supplementary Figure 1a).

#### Drug imputation

In the eICU dataset, approximately one-third of drug names are not recorded across all the 208 hospitals. If these unrecorded drugs are represented as “0”, we tend to have an extremely sparse input, which hampers the training of density estimators due to the lack of informative features. For instance, if density estimators like MADE, VAE, or VQ-VAE are trained under such conditions, these auto-encoders tend to reconstruct more zeros than ones. To address this challenge, we implemented a strategy for imputing the missing drug names. While some drugs lack recorded names, we observed that they possess Hierarchical Ingredient Code List (HICL) codes —– a standardized coding system for identifying healthcare products. Consequently, we could match these unrecorded drugs with their counterparts with both HICL codes and recorded names in the database. This imputation method allows us to fill in missing drug names based on their HICL codes. As a result, the proportion of unrecorded drugs in our dataset decreases to around 20%, which enhances the accuracy and reliability of our models by providing a richer set of features (Figure 1d-e).

#### Drug harmonization

We discovered that different hospitals may use different drug names, although they share the same identity (e.g. “aspirin” and “acetylsalicylic acid”). We also found that the dosage information is included in some drug names (e.g. “aspirin 10 mg”). Consequently, we have minimal overlap in drug names between hospitals, further exacerbating the challenge of sparse input data and hindering the training process. To address this, we implemented a harmonization process to standardize drug names. We merged those with shared identities while disregarding dosage details. Our algorithm comprises two main steps:

- Initial drug mapping: We maintained a reference panel of drug names on the server, containing the 237 most common drugs [97], which is periodically updated and shared with client hospitals. For each drug name from a client hospital (e.g., “aspirin 10mg”), the algorithm checks against each name in the reference panel to find if the reference name (e.g., “aspirin”) is included in the client’s drug name (“aspirin 10mg”). If a match is detected, the client’s drug name is mapped to the reference drug name (e.g., “aspirin”). This approach enables accurate mapping of most drug names without dosage information.
- Use of BioWordVec for mapping: If no direct match is found, the algorithm initiates a similarity analysis. First, the client’s drug name and the reference drug name are converted into their respective word embeddings using BioWordVec [98]. Then, we compute the cosine similarity between these two word embeddings. After comparing with all reference drug names, the reference name with the highest cosine similarity is selected. This approach effectively matches drugs with similar identities.

After harmonization, the proportion of overlapping drugs across hospitals significantly increases (Figure 1f-g).

#### Lab tests data preprocessing

We also included lab tests as model input. However, we encountered several challenges due to discrepancies between the eICU and MIMIC-III datasets. The eICU dataset contains 158 unique lab tests, whilst MIMIC-III includes 590 unique lab tests, with differences in test names, abbreviations, and units across the two datasets. Referring to a previous study that focused on the 29 most influential lab tests for model training [97], we identified 12 lab tests common to both datasets (Supplementary Table 4). Due to variations in the scale of lab results, we normalized the data to ensure consistency and comparability.

#### Time series data preprocessing

We used the eICU dataset to construct predictive models for patients’ mortality, ventilator use, and sepsis diagnosis, and ICU length of stay. To prepare data for mortality and length of stay prediction, we segmented the dataset into two intervals: drug data within the initial 48 hours of ICU admission and mortality outcomes and length of stay after 48 hours. The models were trained using patient demographics and drug administration data from the first 48 hours to predict mortality and length of stay beyond this period. For ventilator use and sepsis diagnosis, we designed a 72-hour observation period after ICU entry, further divided into six 12-hour intervals. The objective was to use drug and demographic information from each interval to predict ventilator use and sepsis occurrence in the next interval.

### Simulation study

#### Generating inputs

We created the simulation dataset input from the preprocessed eICU dataset, which included imputed and harmonized drug data alongside age, sex, BMI, and ethnicity information. For simplicity, we assumed one source hospital and one target hospital in the network. To generate the simulation dataset, we utilized the input from Hospital 167, which has the most patients, as the target input **X**_*τ*_. We further combined the remaining 9 hospitals as the source input **X**_*k*_.

#### Generating labels

We simulated labels by first creating a linear model *f* (**X**) = **Xw** + **b**, where the weight was sampled from Gaussian distribution: **w** ~*𝒩* (0, **I**). Meanwhile, we created a binary mask sampled from Bernoulli distribution: **m** ~ *Bernoulli*(0.15). Then we applied the binary mask **m** to the weight: **w** = **w** ⊙ **m**. As a result, 15% of the weight values were sampled from Gaussian distribution while setting the rest of weight values as 0, which simulated around 15% of features are causal.

In terms of the bias of the linear model **b** ∈ ℝ^*N*^, we set it as: 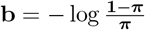 where ***π*** = [0.15]_×*N*_. Then we created imbalanced dataset with most labels were 0, which simulated the real-world scenario.

Finally, we generated simulated labels for source hospital: **y**_*k*_ = 𝕀 [*f* (**X**_*k*_) *>* 0], and target hospital **y**_*τ*_ = 𝕀 [*f* (**X**_*τ*_) *>* 0]. We used the same linear model *f* to generate source and target labels, ensuring consistent conditional probabilities and simulating the covariate-shift problem.

### Experimental design for reliable model evaluation

To enable early stopping and hyperparameter selection, we split the target hospital data into a 50% valiadation set and a 50% test set. The validation set, represented by 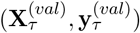, assumes partial label availability at the target hospital. The remaining data from the target hospital, without accessible labels, formed the test set,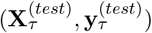.

For a stable assessment of the model performance, our experimental design employed multiple seeds. We selected the 5 hospitals (Hospital 167, 199, 252, 420, 458) from the 10 hospitals with the most patients as the target hospitals. We looped these 5 hospitals, and each hospital under the loop was alternately designated as the target, while the remaining 9 were the source hospitals. Models were trained on the source hospitals and evaluated on the target. We employed bootstrap sampling to generate 100 distinct test sets based on the existing test set 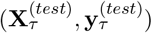. We then evaluated our model on these 100 test sets, obtaining 100 individual results. By calculating the mean and standard deviation of these results, we ensured a more reliable assessment of the model’s performance.

### Hyperparameter selection for FedProx

We compared FedWeight with FedProx [31]. FedProx’s performance was highly sensitive to the choice of its proximal term regularization coefficient (*λ*). While the proximal term is intended to mitigate client drift by constraining local updates, an improperly chosen *λ* can either excessively limit learning or fail to address data heterogeneity—both of which can degrade performance. To ensure a fair and meaningful comparison, we carefully tuned *λ* for each target hospital using its validation data. This per-hospital tuning was performed via grid search to identify the optimal *λ* value that yielded the best performance, ensuring that the reported results for FedProx reflect its optimal performance under well-calibrated conditions.

## Data Availability

All data produced in the present study are available upon reasonable request to the authors.
All data produced in the present work are contained in the manuscript.
All data produced are available online at eICU Collaborative Research Database (https://eicu-crd.mit.edu/about/eicu/) and PhysioNet (http://mimic.physionet.org).

## Data availability

The eICU dataset analyzed in this paper are publicly available through eICU Collaborative Research Database (https://eicu-crd.mit.edu/about/eicu/). The MIMIC-III dataset are publicly available through PhysioNet (http://mimic.physionet.org).

## Code availability

We implemented FedWeight in Python3.9. The software is available at: https://github.com/li-lab-mcgill/FedWeight.

## Acknowledgments

Y.L. is supported by NOVA-FRQNT-NSERC grant (FRQ-NT 2023-NOVA-328677), Canada Research Chair (Tier 2) in Machine Learning for Genomics and Healthcare (CRC-2021-00547) and Natural Sciences and Engineering Research Council (NSERC) Discovery Grant (RGPIN-2016-05174). H.Z. is supported by Artificial Intelligence for Public Health (AI4PH) and Health Equity Trainee Scholarship. D.L. is supported by Singapore NUHS seed fund, (25-0381-A0001)

## Author contributions

Y.L., D.L., and H.Z. conceived the idea. Y.L., D.L., and D.B. supervised the work. H.Z. and J.B. implemented the software and conducted the experiments. Y.L., D.L., D.B., J.B., and H.Z. analyzed the data with the help from N.L.. All authors contributed to the final writing of the manuscript.

## Declaration of Interests

The authors declare no competing interests.

## Notes

### Competing Interest Statement

The authors have declared no competing interest.

### Author Declarations

The study used ONLY openly available patient records that were originally located at: eICU Collaborative Research Database (https://eicu-crd.mit.edu/about/eicu/) and PhysioNet (http://mimic.physionet.org)

### Summary of Updates

Add new experimental results comparing with other FL methods. Reorganized structure of the main text.

