## Supplementary Information for "FedWeight: Mitigating Covariate Shift of Federated Learning on Electronic Health Records Data through Patients Re-weighting"

**Supplementary Figure 1.** Intra-dataset covariate shifts in the MIMIC-III dataset. **a.** K-mean clustering MIMIC-III drugs. **b.** Negative log of p-values from Fisher-exact test for admission types. "NEWBORN" indicates admissions pertaining to patient's birth. "EMERGENCY" indicates unplanned medical care. "ELECTIVE" indicates unplanned admission. **c.** Negative log of p-values from Fisher-exact test for surgery procedures.

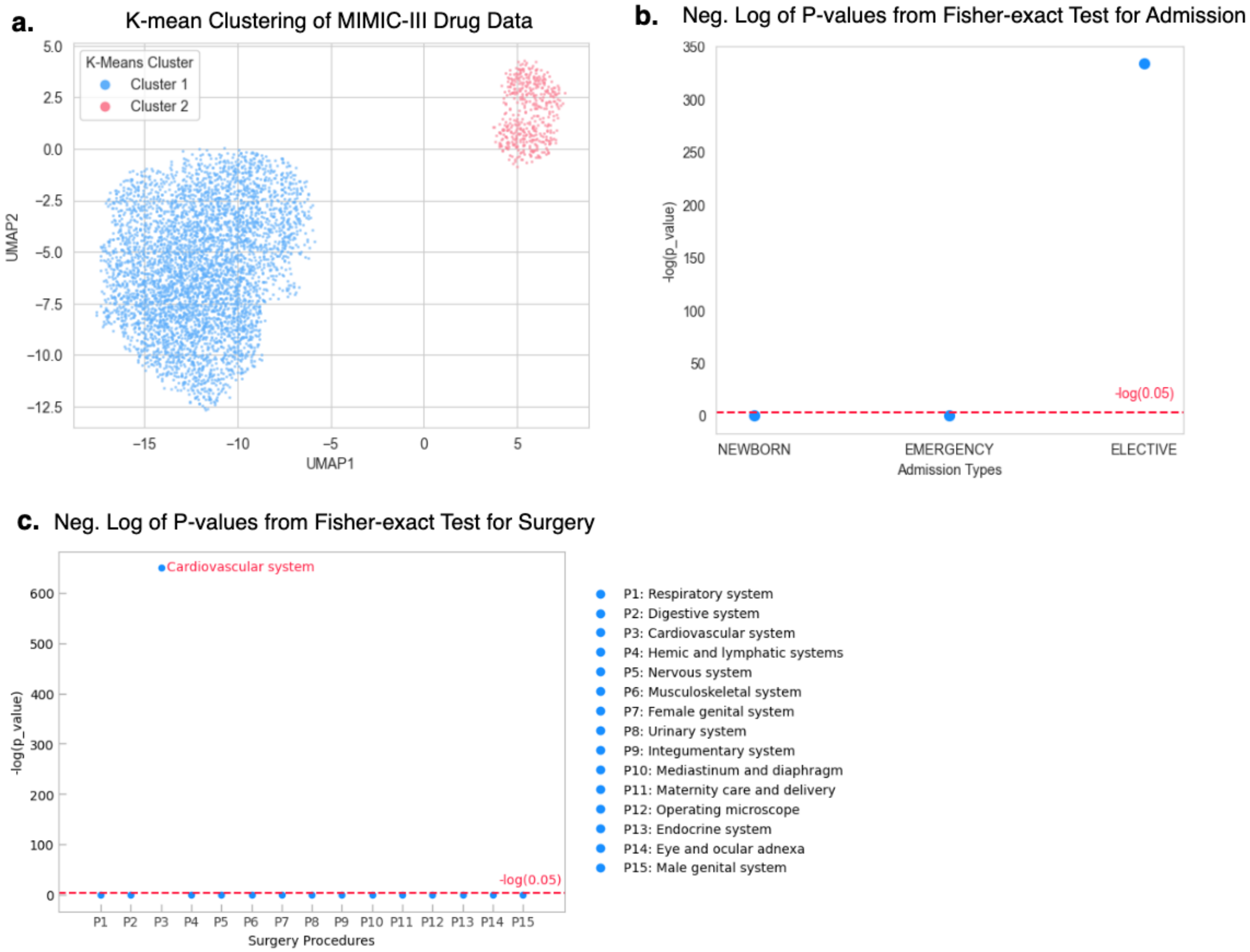

**Supplementary Figure 2.** Performance of FedWeight on the simulation dataset. **a.** AUPRC for model predictions. **b.** AUPRC compared model weights and influential features. **c.** Pearson correlation calculated between the weights of the trained and reference models. We evaluated their statistical significance using the Wilcoxon test against the baseline, denoting * for test p-values < 0.05 and ** for p-values < 0.01.

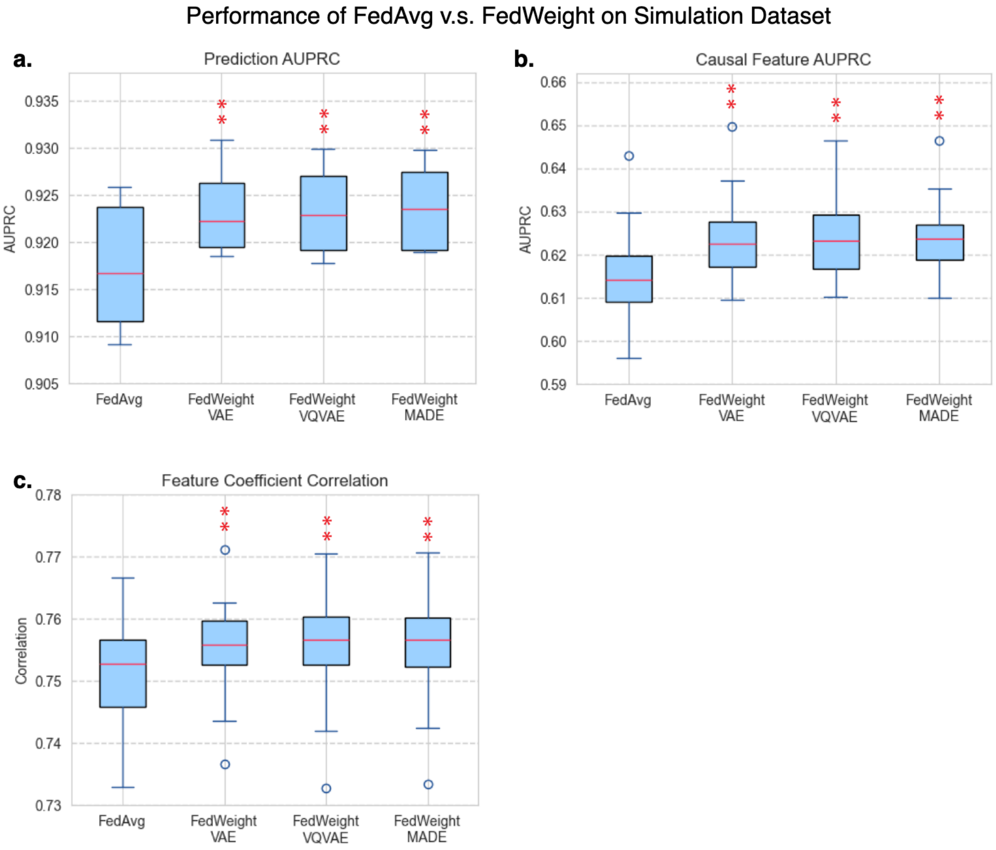

**Supplementary Figure 3.** Sample re-weight correlation analysis across three FedWeight density estimators: MADE, VAE, and VQ-VAE. For this study, we selected two eICU hospitals (420 and 167) with the largest patient populations. Hospital 420 was selected as the source hospital, and Hospital 167 as the target hospital. Using three selected density estimators, we calculated sample re-weights for source hospital patients. Then, we calculated the Pearson correlation between these re-weights generated by MADE, VAE, and VQ-VAE. **a.** Pairwise comparison of sample re-weights assigned by MADE and VAE (Pearson correlation: 0.695). **b.** Pairwise comparison of sample re-weights assigned by MADE and VQ-VAE (Pearson correlation: 0.719). **c.** Pairwise comparison of sample re-weights assigned by VAE and VQ-VAE (Pearson correlation: 0.610).

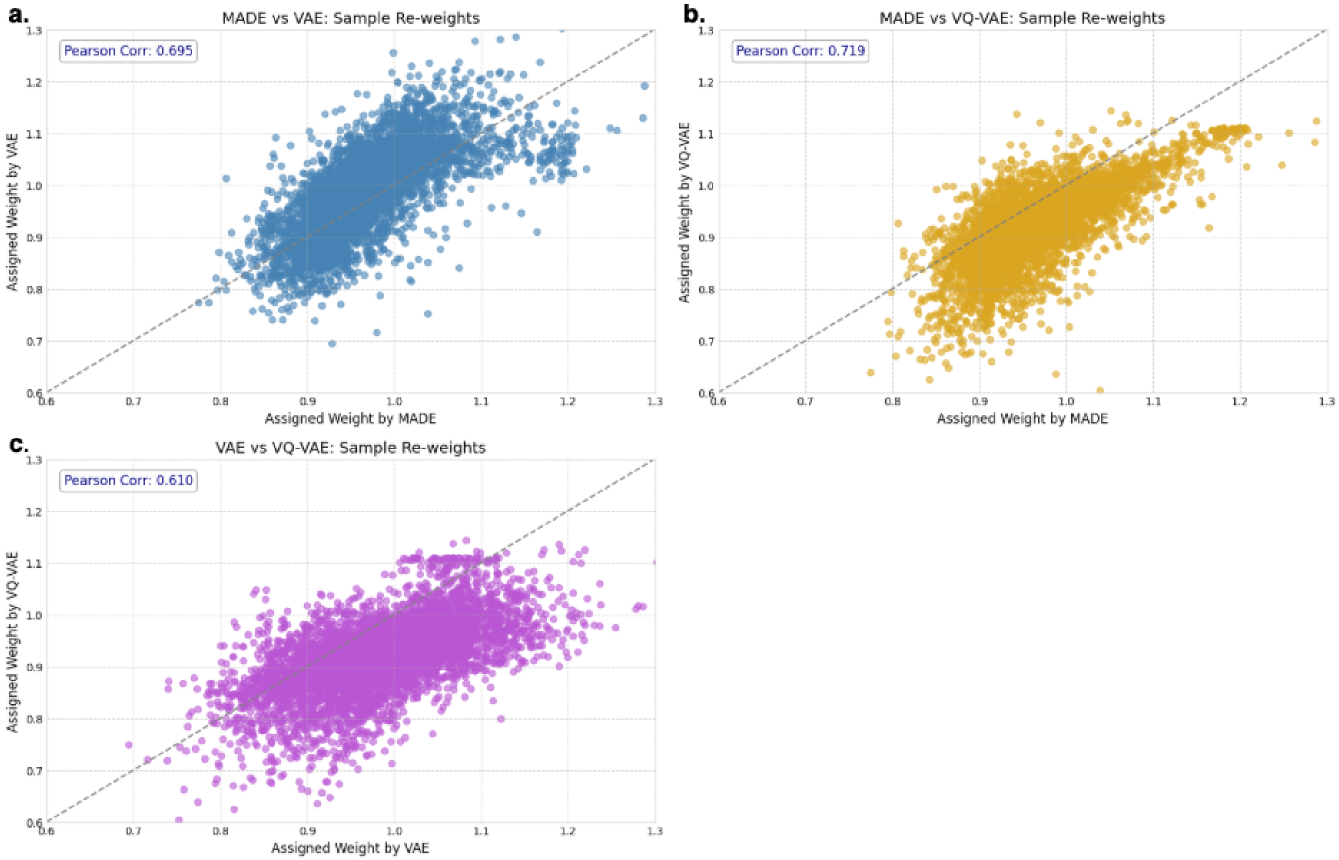

**Supplementary Figure 4.** Performance variation of FedWeightVAE with respect to the number of training epochs of the target density estimator (VAE) across four clinical prediction tasks, evaluated on eICU Hospital 458. The green curve represents the task model's performance (AUPRC for mortality, ventilator, and sepsis prediction; loss for ICU length of stay), while the orange curve denotes the validation likelihood of the VAE. **a.** Mortality prediction. **b.** Ventilator prediction. **c.** Sepsis prediction. **d.** ICU length of stay prediction.

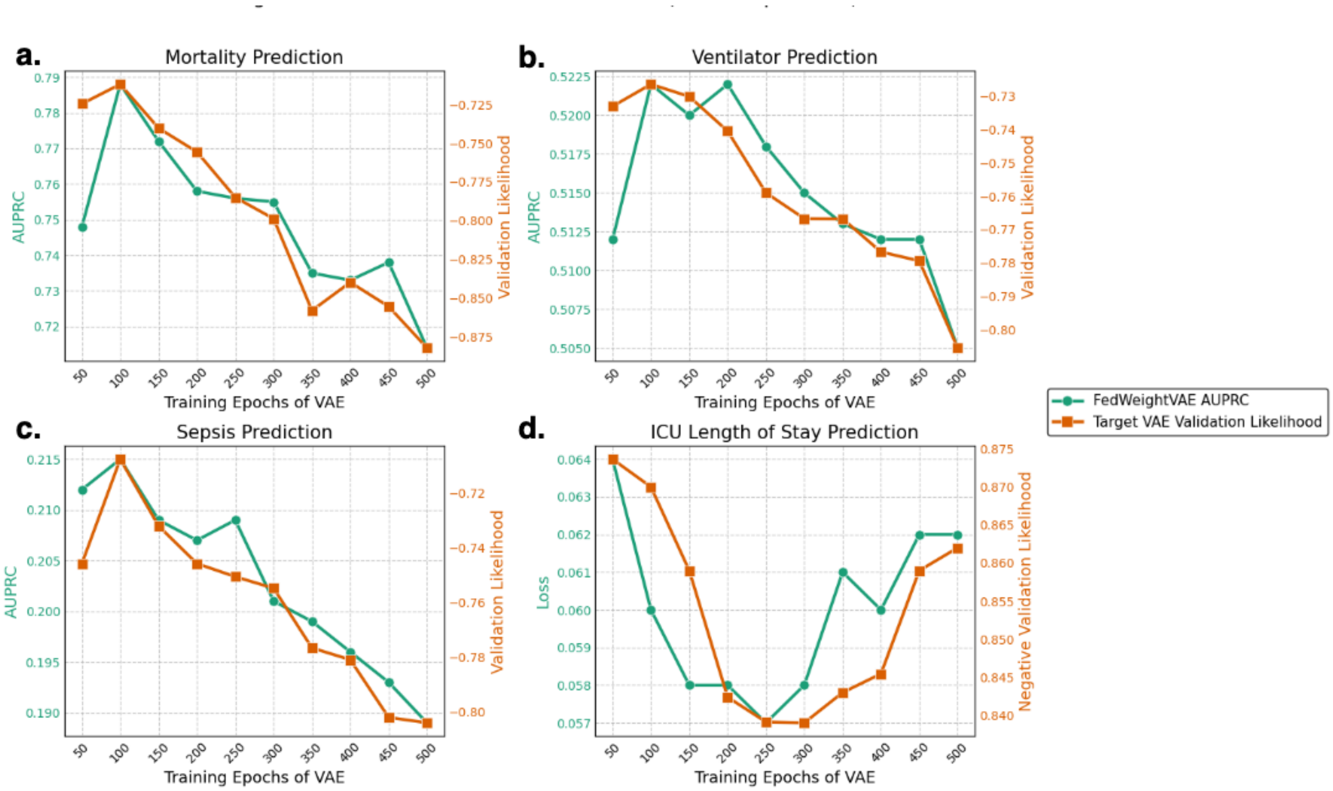

**Supplementary Figure 5.** Performance comparison of ETM-based models in predicting readmission mortality. We used first-admission ICD codes as input to ETM-based models to generate patient-topic mixtures. A logistic regression model was then used to predict readmission mortality based on these mixtures. **a.** Performance on eICU. **b.** Performance on MIMIC-III.

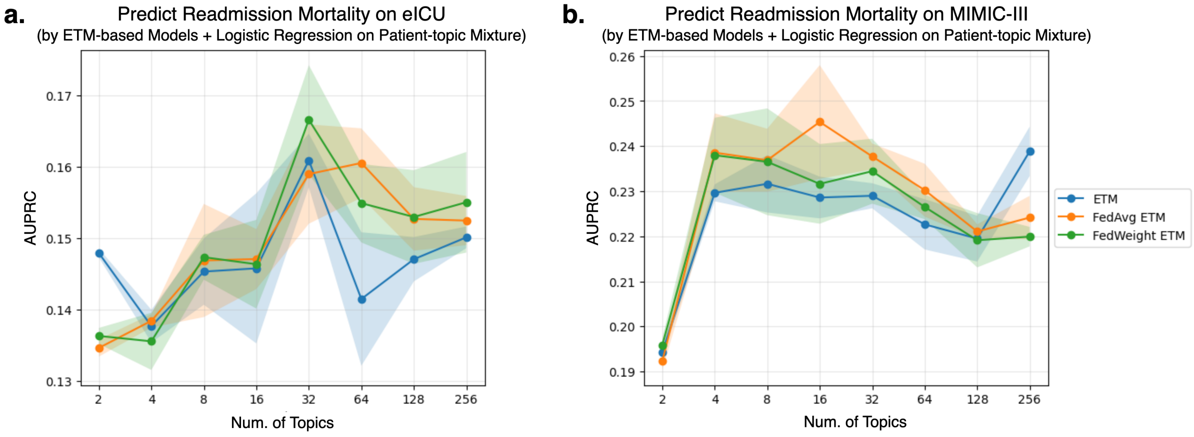

**Supplementary Figure 6.** Percentage of missing values for patient demographics (age, sex, BMI, and ethnicity) and clinical outcomes (mortality, ventilator, sepsis, and ICU length of stay) in the eICU and MIMIC-III datasets. A value is considered missing if it is null or marked as "unknown". Percentage was calculated as the number of missing values divided by the total number of admissions in each dataset.

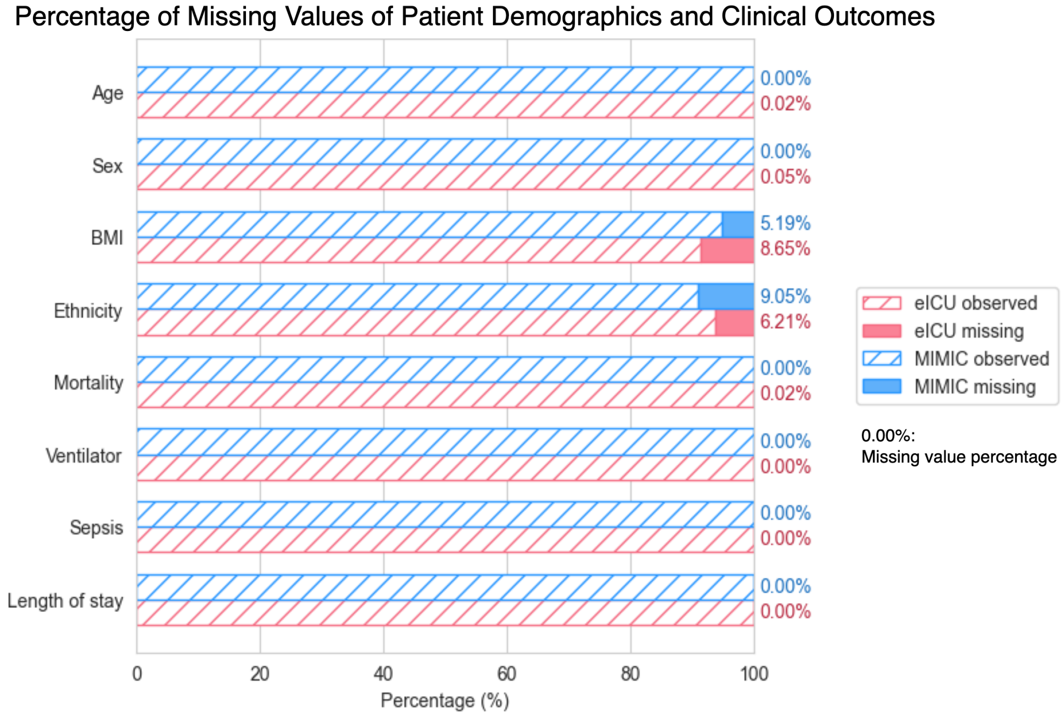

**Supplementary Table 1.** Clinical outcome prediction of standalone and federated methods evaluated on eICU Hospital 167. For the standalone method, we trained separate models on data from each of the nine source hospitals (420, 199, 458, 252, 165, 148, 281, 449, and 283) independently, and each model was subsequently evaluated individually on the target hospital 167. In comparison, for the federated methods, the model was trained collaboratively on the nine source hospitals and evaluated on target hospital 167. For mortality, ventilator, and sepsis prediction, we employed AUPRC as the evaluation metric. We utilised loss for ICU length of stay prediction. Colour **red** was used to highlight the method with the highest AUPRC values and **blue** was applied to denote the method achieving the lowest loss values.

| **Algorithm** | **Source Hospital** | **Mortality** (AUPRC ↑) | **Ventilator** (AUPRC ↑) | **Sepsis** (AUPRC ↑) | **Length of Stay** (Loss ↓) |
|---|---|---|---|---|---|
| Standalone | 420 | 0.546±0.042 | 0.312±0.013 | 0.181±0.013 | 0.070±0.004 |
| | 199 | 0.561±0.045 | 0.420±0.015 | 0.167±0.016 | 0.068±0.003 |
| | 458 | 0.468±0.049 | 0.405±0.013 | 0.145±0.012 | 0.067±0.003 |
| | 252 | 0.493±0.045 | 0.403±0.015 | 0.188±0.019 | 0.069±0.004 |
| | 165 | 0.580±0.046 | 0.487±0.015 | 0.191±0.016 | 0.066±0.003 |
| | 148 | 0.525±0.043 | 0.436±0.013 | 0.208±0.019 | 0.078±0.005 |
| | 281 | 0.463±0.046 | 0.394±0.013 | 0.180±0.016 | 0.070±0.004 |
| | 449 | 0.415±0.045 | 0.343±0.014 | 0.127±0.012 | 0.091±0.006 |
| | 283 | 0.397±0.042 | 0.384±0.013 | 0.203±0.019 | 0.076±0.004 |
| FedAvg | N/A | 0.625±0.045 | 0.514±0.013 | 0.257±0.023 | 0.057±0.004 |
| FedWeightMADE | N/A | 0.630±0.043 | 0.519±0.016 | 0.266±0.022 | **0.056±0.003** |
| FedWeightVAE | N/A | 0.637±0.038 | **0.521±0.015** | 0.283±0.020 | **0.056±0.004** |
| FedWeightVQVAE | N/A | **0.663±0.044** | 0.517±0.013 | **0.284±0.021** | 0.057±0.004 |

**Supplementary Table 2.** Performance comparison of federated models across clinical outcome predictions, including mortality, ventilator, sepsis, and ICU length of stay prediction. FedWeight methods were compared with FedAvg, FedProx, and a centralized model (i.e., model trained on the pooled eICU data and corrected by the covariate indicator variable for each hospital). Performance of clinical outcome predictions on eICU, which was evaluated on a bootstrap-sampled dataset from five target hospitals (167, 199, 252, 420, and 458), with mean and standard deviation computed from the bootstrap samples. Performance of cross-dataset federated model trained on eICU and evaluated on a bootstrapped test set of MIMIC-III. For mortality, ventilator, and sepsis prediction, we employed AUPRC as the evaluation metric. We utilised loss for ICU length of stay prediction. Colour **red** was used to highlight the federated method with the highest AUPRC values, and **blue** was applied to denote the federated method achieving the lowest loss values.

| **Task** | **Algorithm** | **Target Hospital ID** | | | | | **Cross Dataset** |
|---|---|---|---|---|---|---|---|
| | | 167 | 199 | 252 | 420 | 458 | |
| Mortality (AUPRC ↑)) | FedAvg | 0.627±0.044 | 0.630±0.047 | 0.656±0.043 | 0.704±0.027 | 0.741±0.032 | 0.207±0.024 |
| | FedProx | 0.633±0.047 | **0.655±0.039** | 0.642±0.039 | 0.743±0.031 | 0.756±0.036 | **0.235±0.028** |
| | FedWeightMADE | 0.630±0.043 | 0.645±0.042 | 0.662±0.042 | 0.731±0.027 | 0.766±0.032 | 0.214±0.027 |
| | FedWeightVAE | 0.635±0.041 | 0.634±0.042 | **0.676±0.044** | **0.744±0.023** | 0.760±0.030 | 0.216±0.027 |
| | FedWeightVQVAE | **0.668±0.043** | 0.645±0.040 | 0.674±0.040 | 0.728±0.024 | **0.781±0.031** | 0.219±0.023 |
| | Centralized | 0.673±0.044 | 0.654±0.038 | 0.683±0.043 | 0.742±0.026 | 0.754±0.030 | 0.224±0.022 |
| Ventilator (AUPRC ↑)) | FedAvg | 0.515±0.014 | 0.496±0.015 | 0.482±0.016 | 0.427±0.010 | 0.509±0.012 | 0.043±0.005 |
| | FedProx | 0.496±0.013 | 0.488±0.013 | 0.485±0.012 | 0.444±0.011 | 0.505±0.011 | **0.051±0.006** |
| | FedWeightMADE | 0.519±0.017 | 0.500±0.013 | 0.489±0.013 | 0.435±0.011 | 0.516±0.011 | 0.049±0.005 |
| | FedWeightVAE | **0.522±0.018** | 0.514±0.015 | 0.492±0.013 | 0.441±0.012 | **0.522±0.013** | **0.051±0.006** |
| | FedWeightVQVAE | 0.518±0.013 | **0.518±0.013** | **0.503±0.014** | **0.449±0.011** | 0.516±0.010 | 0.048±0.006 |
| | Centralized | 0.531±0.013 | 0.513±0.013 | 0.497±0.013 | 0.451±0.010 | 0.526±0.011 | 0.048±0.006 |
| Sepsis (AUPRC ↑)) | FedAvg | 0.258±0.024 | 0.177±0.019 | 0.267±0.018 | 0.372±0.017 | 0.174±0.027 | 0.033±0.010 |
| | FedProx | 0.264±0.023 | **0.198±0.019** | 0.272±0.021 | **0.389±0.018** | 0.201±0.035 | **0.063±0.031** |
| | FedWeightMADE | 0.266±0.024 | 0.185±0.017 | **0.282±0.024** | 0.380±0.017 | 0.192±0.026 | 0.038±0.016 |
| | FedWeightVAE | 0.284±0.020 | 0.184±0.021 | 0.277±0.021 | 0.384±0.018 | **0.211±0.036** | 0.040±0.011 |
| | FedWeightVQVAE | **0.285±0.021** | 0.179±0.018 | 0.267±0.020 | 0.385±0.018 | 0.209±0.037 | 0.037±0.014 |
| | Centralized | 0.280±0.017 | 0.183±0.018 | 0.270±0.021 | 0.382±0.015 | 0.197±0.031 | 0.046±0.017 |
| Length of Stay (Loss ↓)) | FedAvg | 0.057±0.004 | 0.050±0.003 | 0.052±0.004 | 0.054±0.002 | 0.061±0.004 | 0.132±0.007 |
| | FedProx | 0.060±0.004 | 0.049±0.003 | **0.050±0.003** | 0.054±0.003 | **0.059±0.004** | **0.108±0.005** |
| | FedWeightMADE | **0.056±0.004** | 0.048±0.002 | 0.051±0.003 | 0.054±0.002 | 0.060±0.004 | 0.121±0.007 |
| | FedWeightVAE | **0.056±0.004** | 0.048±0.003 | **0.050±0.003** | 0.054±0.002 | 0.060±0.004 | 0.119±0.006 |
| | FedWeightVQVAE | 0.057±0.004 | **0.046±0.003** | 0.051±0.003 | **0.053±0.002** | 0.060±0.003 | 0.111±0.006 |
| | Centralized | 0.055±0.003 | 0.048±0.003 | 0.049±0.002 | 0.054±0.002 | 0.059±0.004 | 0.109±0.006 |

**Supplementary Table 3.** Notations in FedWeight

| Notation | Description |
|---|---|
| $K$ | Total number of source hospitals |
| $\tau$ | Target hospital in the federated network |
| $k$ | Source hospital in the federated network |
| $D$ | Feature size across the federated network |
| $N_k$ | Number of patients in source hospital $k$ |
| $N$ | Total number of patients across all source hospitals |
| $\mathbf{X}$ | Input data for hospital |
| $\mathbf{y}$ | Labels for hospital |
| $p$ | Density estimator for hospital |
| $f$ | The linear model used in generating simulation labels |
| $\mathbf{w}_k$ | Local model parameters for source hospital $k$ |
| $\mathbf{w}$ | Aggregated model parameters for target hospital $\tau$ |
| $\mathbf{q}$ | Model parameters for the ETM encoder |
| $\mathbf{u}$ | Model parameters for the ETM linear layer which outputs $\boldsymbol{\mu}$ |
| $\mathbf{s}$ | Model parameters for the ETM linear layer which outputs $\boldsymbol{\sigma}$ |
| $\boldsymbol{\rho}$ | ETM ICD embedding |
| $\boldsymbol{\alpha}$ | ETM topic embedding |
| $\boldsymbol{\theta}$ | ETM patient-topic mixture |
| $\boldsymbol{\beta}$ | ETM topic-ICD mixture |
| $\mathbf{Z}$ | ETM latent representation |
| $\varphi$ | The re-weighting ratio used in the weighted log-likelihood algorithm |
| $\lambda$ | Hyper-parameter that controls the degree of re-weighting |

**Supplementary Table 4.** Lab tests, abbreviations, and units

| Lab Tests | Abbreviations | Units |
|---|---|---|
| Oxygen Saturation | o2sat | % |
| Partial pressure of oxygen | pao2 | mm Hg |
| Partial pressure of carbon dioxide | paco2 | mm Hg |
| pH value of blood | ph | Units |
| Albumin | albumin | g/dL |
| Bands | bands | % |
| Blood urea nitrogen | bun | mg/dL |
| Hematocrit | hct | % |
| International Normalized Ratio (PT test) | inr | ratio |
| Lactate | lactate | mmol/L |
| Platelets | platelets | K/mcL |
| White blood cell | wbc | K/mcL |

**Supplementary Note 1. Convergence analysis of FedWeight under covariate shifts**

**Problem definition and notations**

In FedWeight, we consider the federated optimization problem under covariate shift. Suppose there are $K$ source hospitals indexed by $k$. The global objective of FedWeight is defined as:

$$F^{\text{FedWeight}}(\mathbf{w}) = \sum_{k=1}^{K} \frac{N_k}{N} \mathbb{E}_{\mathbf{x}\sim p_k(\mathbf{x})}[\varphi_k(\mathbf{x})\ell(\mathbf{w};\mathbf{x})],$$

where $N_k$ is the number of patients in hospital $k$, $N$ denotes the total number of patients, $p_k(\mathbf{x})$ represents the source data distribution at hospital $k$, $\varphi_k(\mathbf{x}) = \left(\frac{p_\tau(\mathbf{x})}{p_k(\mathbf{x})}\right)^\lambda$ is the re-weight in FedWeight, $p_\tau(\mathbf{x})$ represents the data distribution of the target hospital, and $\ell(\mathbf{w};\mathbf{x}) = \log p(y|\mathbf{x}, \mathbf{w}_k)$.

We also define the local weighted objective as:

$$F_k^{\text{FedWeight}}(\mathbf{w}) = \mathbb{E}_{\mathbf{x}\sim p_k(\mathbf{x})}[\varphi_k(\mathbf{x})\ell(\mathbf{w};\mathbf{x})].$$

Thus:

$$F^{\text{FedWeight}}(\mathbf{w}) = \sum_{k=1}^{K} \frac{N_k}{N} F_k^{\text{FedWeight}}(\mathbf{w}).$$

Let $\mathbf{w}^*$ be the global minimizer:

$$\mathbf{w}^* = arg\min_{\mathbf{w}} F^{FedWeight}(\mathbf{w}).$$

**Assumptions**

Following the convergence analysis framework used in FedAvg [1], we establish the following four standard assumptions on each local weighted objective function $F_k^{\text{FedWeight}}(\mathbf{w})$:

**Assumption 1** ($L$-Smoothness): Each local objective function is assumed to be $L$-smooth, i.e., for any $\mathbf{u}, \mathbf{v}$, we have:

$$F_k^{\text{FedWeight}}(\mathbf{u}) \leq F_k^{\text{FedWeight}}(\mathbf{v}) + \nabla F_k^{\text{FedWeight}}(\mathbf{v})^\top(\mathbf{u}-\mathbf{v}) + \frac{L}{2}\|\mathbf{u}-\mathbf{v}\|^2.$$

**Assumption 2** ($\mu$-Strong Convexity): Each local objective is $\mu$-strongly convex, i.e., for any $\mathbf{u}, \mathbf{v}$, we have:

$$F_k^{\text{FedWeight}}(\mathbf{u}) \geq F_k^{\text{FedWeight}}(\mathbf{v}) + \nabla F_k^{\text{FedWeight}}(\mathbf{v})^\top(\mathbf{u}-\mathbf{v}) + \frac{\mu}{2}\|\mathbf{u}-\mathbf{v}\|^2.$$

**Assumption 3** (Bounded Variance): The stochastic gradient variance of $F_k^{\text{FedWeight}}(\mathbf{w})$ is bounded. Specifically, there exists a constant $\sigma_k^2$ and $C_\varphi$ satisfying:

$$\mathbb{E}\|\nabla F_k^{\text{FedWeight}}(\mathbf{w};\xi) - \nabla F_k^{\text{FedWeight}}(\mathbf{w})\|^2 \leq \sigma_k^2 C_\varphi,$$

where the variance-increase constant $C_\varphi$ is defined by:

$$C_\varphi = \mathbb{E}_{\mathbf{x}\sim p_k(\mathbf{x})}[\varphi_k(\mathbf{x})^2] < \infty.$$

**Assumption 4** (Bounded Gradient Norm): The squared norm of the stochastic gradient is bounded, i.e., there exists constant $G$ such that:

$$\mathbb{E}\|\nabla F_k^{\text{FedWeight}}(\mathbf{w};\xi)\|^2 \le G^2 C_\varphi.$$

**Convergence analysis**

Let $\mathbf{w}_t$ be the global model at communication round $t$. At the start, each hospital $k$ initializes its local model as:

$$\mathbf{w}_t^{k,0} = \mathbf{w}_t,$$

and performs $E$ local SGD updates with learning rate $\eta_t$ on its reweighted loss $F_k^{FedWeight}$:

$$\mathbf{w}_t^{k,i+1} = \mathbf{w}_t^{k,i} - \eta_t \nabla F_k^{\text{FedWeight}}(\mathbf{w}_t^{k,i};\xi_t^{k,i}), \quad i = 0,\ldots,E-1.$$

Then after $E$ steps, global aggregation at the target hospital side is:

$$\mathbf{w}_{t+1} = \sum_{k=1}^{K} \frac{N_k}{N} \mathbf{w}_t^{k,E}.$$

Define average local updates deviation from global updates:

$$\bar{\mathbf{w}}_{t+1} = \mathbf{w}_t - \eta_t \sum_{k=1}^{K} \frac{N_k}{N} \sum_{i=0}^{E-1} \nabla F_k^{\mathrm{FedWeight}}(\mathbf{w}_t^{k,i};\xi_t^{k,i}),$$

Then we have:

$$\mathbf{w}_{t+1} - \bar{\mathbf{w}}_{t+1} = \eta_t \sum_{k=1}^{K} \frac{N_k}{N} \sum_{i=0}^{E-1} \left[\nabla F_k^{\mathrm{FedWeight}}(\mathbf{w}_t^{k,i};\xi_t^{k,i}) - \nabla F_k^{\mathrm{FedWeight}}(\mathbf{w}_t^{k,i})\right].$$

Taking expectation conditioned on $\mathbf{w_t}$:

$$\mathbb{E}\left[\mathbf{w}_{t+1} - \bar{\mathbf{w}}_{t+1} \mid \mathbf{w}_t\right] = 0.$$

Thus, the update bias is zero-mean. Base on **Assumption 3** and **Assumption 4**, we have:

$$\begin{aligned}\mathbb{E}\left\|\mathbf{w}_{t+1} - \bar{\mathbf{w}}_{t+1}\right\|^2 &= \eta_t^2 \sum_{k=1}^{K} \left(\frac{N_k}{N}\right)^2 \sum_{i=0}^{E-1} \mathbb{E}\left\|\nabla F_k^{\text{FedWeight}}(\mathbf{w}_t^{k,i};\xi_t^{k,i}) - \nabla F_k^{\text{FedWeight}}(\mathbf{w}_t^{k,i})\right\|^2 \\ &\le E\eta_t^2 \sum_{k=1}^{N} \left(\frac{N_k}{N}\right)^2 \sigma_k^2 C_\varphi.\end{aligned}$$

Considering smoothness and strongly convex assumptions (**Assumption 1** and **Assumption 2**) on global objective $F^{FedWeight}$, we have the standard descent lemma:

$$\mathbb{E}\left[F^{\text{FedWeight}}(\mathbf{w}_{t+1})\right] \leq F^{\text{FedWeight}}(\mathbf{w}_t) + \mathbb{E}\left\langle \nabla F^{\text{FedWeight}}(\mathbf{w}_t), \mathbf{w}_{t+1} - w_t \right\rangle + \frac{L}{2}\mathbb{E}\left\|\mathbf{w}_{t+1} - \mathbf{w}_t\right\|^2.$$

Substitue $\mathbf{w_{t+1}}$ into above and taking expectation:

$$\begin{aligned}
\mathbb{E}\left[F^{\text{FedWeight}}(\mathbf{w}_{t+1})\right] - F^{\text{FedWeight}}(\mathbf{w}_t) &\leq -\eta_t \sum_{k=1}^{K}\frac{N_k}{N}\sum_{i=0}^{E-1}\mathbb{E}\left\|\nabla F_k^{\text{FedWeight}}(\mathbf{w}_t^{k,i})\right\|^2 + \frac{L}{2}\mathbb{E}\left\|\mathbf{w}_{t+1} - \mathbf{w}_t\right\|^2 \\
&\leq -\eta_t E\mathbb{E}\left\|\nabla F^{\text{FedWeight}}(\mathbf{w}_t)\right\|^2 \\
&\quad + \frac{L}{2}\left(2E^2\eta_t^2 G^2 C_\varphi + 2E\eta_t^2\sum_{k=1}^{K}\left(\frac{N_k}{N}\right)^2\sigma_k^2 C_\varphi\right).
\end{aligned}$$

After applying strong convexity (**Assumption 2**), we obtain the standard inequality for strongly convex objectives:

$$\mathbb{E}\left[F^{\text{FedWeight}}(\mathbf{w}_{t+1})\right] - F^* \leq (1-\mu E\eta_t)\left[F^{\text{FedWeight}}(\mathbf{w}_t) - F^*\right] + \frac{L\eta_t^2 E}{2}\left[2EG^2C_\varphi + 2\sum_{k=1}^{K}(\frac{N_k}{N})^2\sigma_k^2 C_\varphi\right],$$

where $F^* = \min_{\mathbf{w}} F^{FedWeight}(\mathbf{w})$. We simplify to obtain clearly:

$$\mathbb{E}\left[F^{\text{FedWeight}}(\mathbf{w}_{t+1})\right] - F^* \leq (1-\mu E\eta_t)\left[F^{\text{FedWeight}}(\mathbf{w}_t) - F^*\right] + \eta_t^2 E^2 L G^2 C_\varphi + L\eta_t^2 E\sum_{k=1}^{K}(\frac{N_k}{N})^2\sigma_k^2 C_\varphi.$$

Choosing the learning rate as:

$$\eta_t = \frac{2}{\mu(\gamma+t)}, \quad \text{where} \quad \gamma = \max\left(\frac{8L}{\mu}, E\right),$$

and applying telescoping sums across all $T$ rounds, we have:

$$\mathbb{E}\left[F^{\text{FedWeight}}(\mathbf{w}_T)\right] - F^* \leq \frac{\gamma}{\gamma+T-1}\left[F^{\text{FedWeight}}(\mathbf{w}_1) - F^*\right] + \frac{4\left(E^2G^2C_\varphi + \sum_{k=1}^{K}(\frac{N_k}{N})^2\sigma_k^2 C_\varphi\right)L}{\mu^2(\gamma+T-1)}.$$

Define clearly:

$$B^{\text{FedWeight}} = E^2G^2C_\varphi + \sum_{k=1}^{K}(\frac{N_k}{N})^2\sigma_k^2 C_\varphi.$$

Finally, we obtain the final convergence bound of FedWeight explicitly:

$$\mathbb{E}[F^{\text{FedWeight}}(\mathbf{w}_T)] - F^* \leq \frac{2L/\mu}{\gamma+T-1}\left(\frac{2B^{\text{FedWeight}}}{\mu} + \frac{\mu\gamma}{2}\|\mathbf{w}_1 - \mathbf{w}^*\|^2\right).$$

Thus, FedWeight achieves a clear and rigorous convergence rate of $O(1/T)$.

**Conclusion**

This rigorous convergence analysis establishes that FedWeight, under covariate shift, achieves a convergence rate of $O(1/T)$—matching that of FedAvg [1]. However, FedWeight demonstrates superior performance by explicitly reweighting source hospitals' distributions, ensuring better alignment with the target hospital's data distribution.

### Supplementary Note 2. Model architecture

For the simulation dataset, our objectives are to evaluate the model's predictive performance and interpretability. Due to the ease of linear models in revealing influential features, we opted for a linear model in our simulation studies. In contrast, the real-world mortality and ICU length of stay prediction using the eICU dataset involves leveraging patient demographics and drug data from the initial 48 hours of ICU admission to predict mortality after this period. Therefore, we selected a Multi-Layer Perceptron (MLP) architecture, reflecting its capacity for handling complex, non-linear relationships in high-dimensional data. For predicting ventilator use and sepsis diagnosis, which requires analyzing current interval data to predict outcomes in the next interval, we employed a Long Short-Term Memory (LSTM) architecture [2], owing to its proficiency in capturing temporal dependencies within sequence data.

Regarding our FedWeight density estimators, we utilized a standard MADE architecture. For density estimation with VAE, we employed a beta-VAE with KL divergence annealing to mitigate posterior collapse [3, 4]. In employing VQ-VAE as a density estimator, we adopted an Exponential Moving Average (EMA) method for stable codebook updates [5].
